# Magnetocardiography for the diagnosis of coronary artery disease: a systematic review and meta-analysis

**DOI:** 10.1101/2024.01.31.24302044

**Authors:** Miao Yang, Chenxi Sun, Biying Zhao, Bo Wu, Jing Xiang, Man Xu, Teng Wu, Jianwei Zhang, Wei Xu, Hong Guo

## Abstract

**Background:** Globally, coronary heart disease (CHD) is a major public health issue because it significantly increases mortality and medical expenses. In the recent years, magnetocardiography (MCG) has shown its potential as a new tool for diagnosing CHD. However, the quantitative assessment of MCG currently used for CHD diagnosis are insufficient, preventing its full integration into routine clinical practice.

**Methods:** We searched PubMed (including MEDLINE), Embase, Web of Science, Cochrane Library, Scopus, China National Knowledge Infrastructure (CNKI), Wanfang Data, and ClinicalTrials.gov for studies published up to March 4, 2023. We systematically searched for studies that compare MCG (as the index test) with coronary angiography (as the standard reference) for diagnosing CHD in suspected population. Three authors assessed the quality of the included studies independently using the QUADAS-2 tool. We calculated the pooled value of the sensitivity and specificity of MCG using the bivariate model. To investigate clinical and methodological factors that might contribute to the statistical heterogeneity, we used meta-regression and subgroup analysis. This systematic review and meta-analysis has been conformed to PRISMA guidelines and registered with PROSPERO (number CRD42022332272).

**Results:** By searching, we found 174 studies, 18 of them included 2,571 subjects from 6 countries and regions and met the inclusion criteria. The combined values for sensitivity and specificity are 86% (95% CI: 80-91) and 79% (95% CI: 71-86), respectively. The summary receiver operating characteristic (SROC) curve’s area under the curve was 0.90. We found significant statistical heterogeneity between studies (for sensitivity, I^2^=94.22% (95% CI: 92.48-95.96), Q=294.11, *P*<0.01; for specificity, I^2^=88.98% (95% CI: 84.95-93.01), Q=154.26, *P*<0.01.

**Conclusion:** Given its high sensitivity, MCG has a high value for diagnosing CHD, especially in primary screening. Further investigation is required to examine additional factors that may affect the performance of MCG considering the limited clinical trials and sample size.

## Introduction

Ischemic heart disease (IHD) is of high incidence and mortality rate worldwide, with an estimated 240 million cases globally in 2020.^1^ As the leading cause of death across low-, middle-, and high-income countries,^2^ IHD has a rising percentage of the relevant death continuously.^3^

Coronary heart disease (CHD), the primary cause of IHD, is brought on by the narrowing of coronary arteries and has resulted in a significant medical burden globally. Adults in the United States experienced over 20 million cases of CHD between 2017 and 2020, or about 7.1% of the population.^1^ In Canada, the estimated average annual hospitalization costs for CHD patients are projected to reach 2.2 billion Canadian dollars, not to mention additional economic burdens from medication expenses and loss of productivity.^4^

In recent years, magnetocardiography (MCG) has emerged as a promising method for diagnosing cardiovascular diseases. The cardiac magnetic field generated by the electrical activity of the heart is extremely weak, with a magnitude of about 10^-11^ T. MCG can be accurately measured with a high signal-to-noise ratio using highly sensitive quantum magnetometers positioned above the chest. Since myocardial cells’ electrical activity is what causes the heart to contract, MCG offers a clear reflection of that activity and enables the assessment of cardiac function. Compared to morphological methods such as coronary angiogram (CAG) and echocardiogram (ECHO), MCG identifies cardiac abnormalities before significant morphological changes become apparent, potentially indicating an earlier indication of the presence of lesions. MCG has a higher sensitivity for currents produced by abnormal cardiac activities despite having similar waveforms and origins to electrocardiography (ECG).^5^ Aside from that, MCG might be more sensitive to the subtle signals associated with early-stage disease due to the homogenous magnetic permeability in various tissues.

Multiple studies have also revealed the particular advantages of MCG in detecting CHD. MCG has been shown to identify patients without abnormality of resting ECG,^6^ and has exhibited superior diagnostic performance when compared with ECHO and biochemical parameters such as cardiac troponin.^7^ In addition, recent research has identified a range of potential diagnostic markers for CHD, including QT dispersion,^8^ QRS complex duration,^9^ ST segment-related parameters,^10^ and so on, which hold promise for further enhancing the diagnostic capabilities of MCG.

It is crucial to obtain an accurate assessment of MCG’s diagnostic capacity for the future, especially when considering the availability of flexible, affordable MCG devices based on OPM greatly which increases the viability of its clinical application. Initial clinical trials have been carried out internationally to achieve this. However, a single trial’s sample size is always constrained by the lengthy duration and the challenge of locating suitable samples. It is challenging to come to an unbiased conclusion about the applicability and diagnostic efficacy of MCG because the outcomes of clinical trials can vary significantly due to individual differences, especially when the sample size is insufficient. Additionally, the transition of MCG from a research tool to its clinical application is hampered by the dearth of clinical evidence brought on by these limitations.

Systematic review and meta-analysis are statistical methods that integrate comprehensive results from multiple studies, thereby increasing the sample size and enhancing the accuracy of conclusions. These methods overcome the limitations of individual MCG trials and can provide more reliable evidence for clinical decision-making. Due to the enhanced focus and studies on MCG, adequate clinical trials of MCG can be integrated together for accurate evaluation as well as further conclusions. At the same time, MCG exhibits variations across diverse demographic groups, such as those with different ages, genders, and disease severities.^11^ The MCG itself also has different measurement modes and diagnostic indicators.^12^ A detailed analysis is necessary to identify the specific populations and circumstances where MCG is most effective, and provide more tailored guidance for clinical decision-making.

## Methods

### Search strategy and selection criteria

We reported findings based on the PRISMA guidelines^13^ (details in supplementary materials), and a review protocol was registered in PROSPERO (CRD42022332272). For this systematic review and meta-analysis, we searched PubMed (including MEDLINE), Embase, Web of Science, Cochrane Library, Scopus, China National Knowledge Infrastructure (CNKI), Wanfang Data, and ClinicalTrials.gov for studies published up to March 4, 2023. We included all studies published in any language about patients with suspected CHD, which report the diagnostic results of MCG (as index test) and CAG (as reference standard) in all subjects. Studies based on immature machine learning are excluded due to applicability concerns. Studies should be cross-sectional design. To be eligible, studies should recruit patients with suspected CHD symptoms, and all subjects should undergo both MCG and CAG, and we can obtain the value of the entries in the diagnostic 2×2 table through the information provided in the article (details in supplementary materials).

### Data extraction

CS, BZ and MY independently screened the titles and abstracts of the articles obtained from the database searches to initially exclude studies against eligibility criteria. They then conducted a thorough examination of the full text of the remaining studies to determine whether they met the inclusion criteria. When dealing with studies containing duplicated data, our approach was to selectively include them based on the principle of avoiding data overlap and ensuring the broadest possible range of patient inclusion times. Any disagreements regarding the inclusion of a study were resolved by WX. Additionally, BZ and CS reviewed the reference lists of the searched studies for potentially relevant articles that were missed during the initial screening process. To ensure accuracy and completeness of the data, BZ and MY contacted relevant researchers to inquire about unpublished trials. BZ and MY extracted the data from eligible studies independently, and the results were verified by CS and WX.

We extracted the following data from the eligible articles: first author, year of publication, subjects, selected diagnostic criteria of CAG and MCG, diagnostic 2×2 table of MCG. If available, we also extracted the diagnostic 2×2 table of ECG. In case where the results of multiple diagnostic indicators for MCG were reported, we extracted the results with the highest Youden Index for further analysis.

### Quality assessment

BZ and MY independently assessed the quality of the included studies based on QUADAS-2 to evaluate the risk of bias and applicability from four aspects, including patient selection, index test, reference standard, and flow and timing. For cases of disagreement, consensus was achieved from CS. The quality assessment data were stored in Microsoft Excel® LTSC MSO (16.0.14332.20540).

### Data analysis

We used the 2×2 table to evaluate the diagnostic performance of MCG (table S11). With STATAMP 18, we employed the bivariate model to the sensitivity and specificity with their corresponding 95% confidence intervals (CI) for each study and the pooled data. Furthermore, we generated a SROC curve to obtain a quantitative evaluation of the diagnostic value of MCG by calculating the area under the SROC curve (AUC).

We utilized Higgins and Thompson’s *I*-squared (*I*^2^) and Cochran’s *Q* to quantitatively assess the degree of statistical heterogeneity among studies. For the meta-regression and subgroup analysis, we explore three potential variables that may result in significant heterogeneity, including the presence of stress tests, the threshold of the gold standard, and the patient’s resting ECG status. The stress test group comprises studies that measured during exercise, recovery from exercise, or after administration of dobutamine, while the resting group only includes studies that measured the MCG of the subjects at rest. The mild group includes studies using a threshold of 50% for CAG, which means that patients with >50% stenosis are diagnosed as CHD, while the severe group use a threshold of 70%. The silent group includes studies that screened suspected CHD patients with normal resting ECG, while the ensemble group included all patients with suspected CHD.

### Role of the funding source

The study design, data collection, data analysis, data interpretation, and manuscript writing were performed without any involvement from the funders. The corresponding author had complete access to all the data in the study and were responsible for the decision to submit for publication.

## Results

From database searches, we found 130 studies. From international trial registries search, we found 44 relevant articles. After removing duplicates and after initial screening, that number dropped to 30. 18 studies were chosen for the meta-analysis after reading the full texts (figure 1). Studies were excluded from this process mainly due to the irrelevant content and various experimental techniques (table S12, table S13).

**Figure 1.**
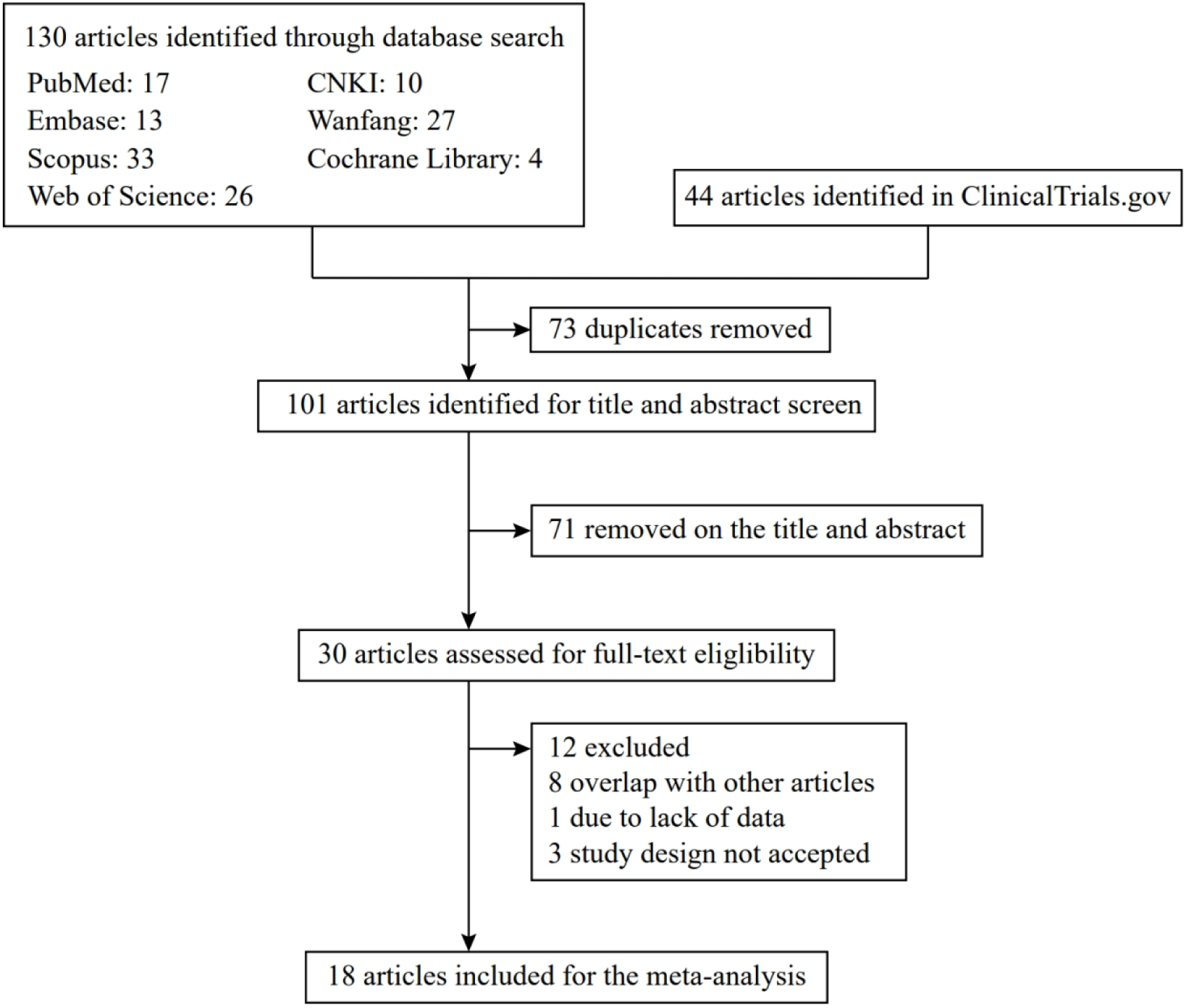
Results of study inclusion.

The 18 eligible studies were published between 2003 and 2023 and included a total of 2,571 participants from 6 different countries and regions: United States,^14,15^ China,^16–22^ Chinese Taiwan,^23,24^ Germany,^7,25–29^ Japan,^30^ and Ukraine^27^ (table 1). 4 studies exclusively recruited suspected CHD patients with normal rest ECG, while the remaining 14 studies included suspected CHD patients regardless of their rest ECG status. 7 studies used a threshold of 50% for diagnosing CHD by CAG, while 10 studies used a threshold of 70%. 4 studies measured stress MCG, with 1 study performing measurements after the administration of dobutamine and 3 studies measuring MCG during or after exercise. 14 studies only measured the rest MCG.

**Table 1.**
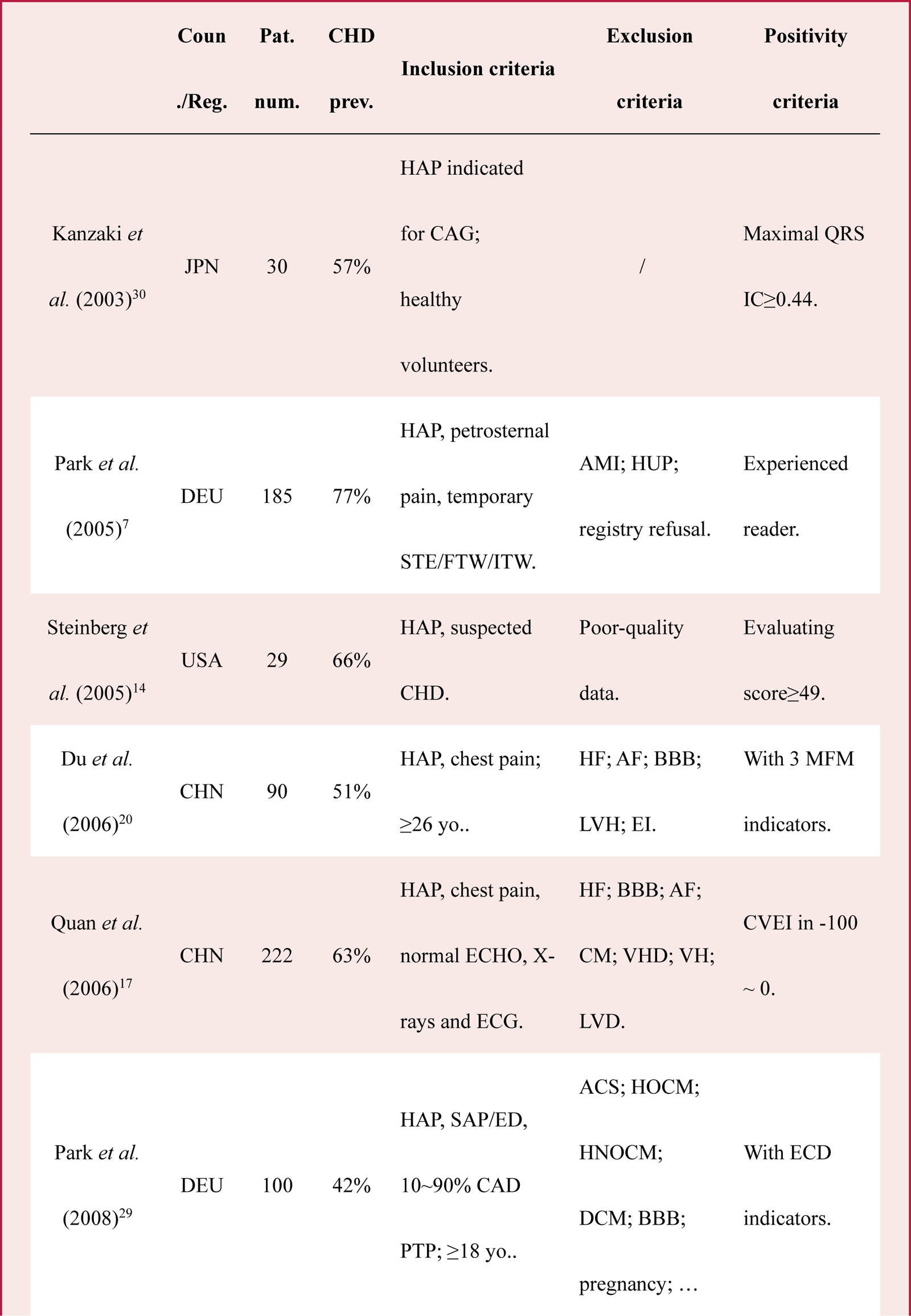
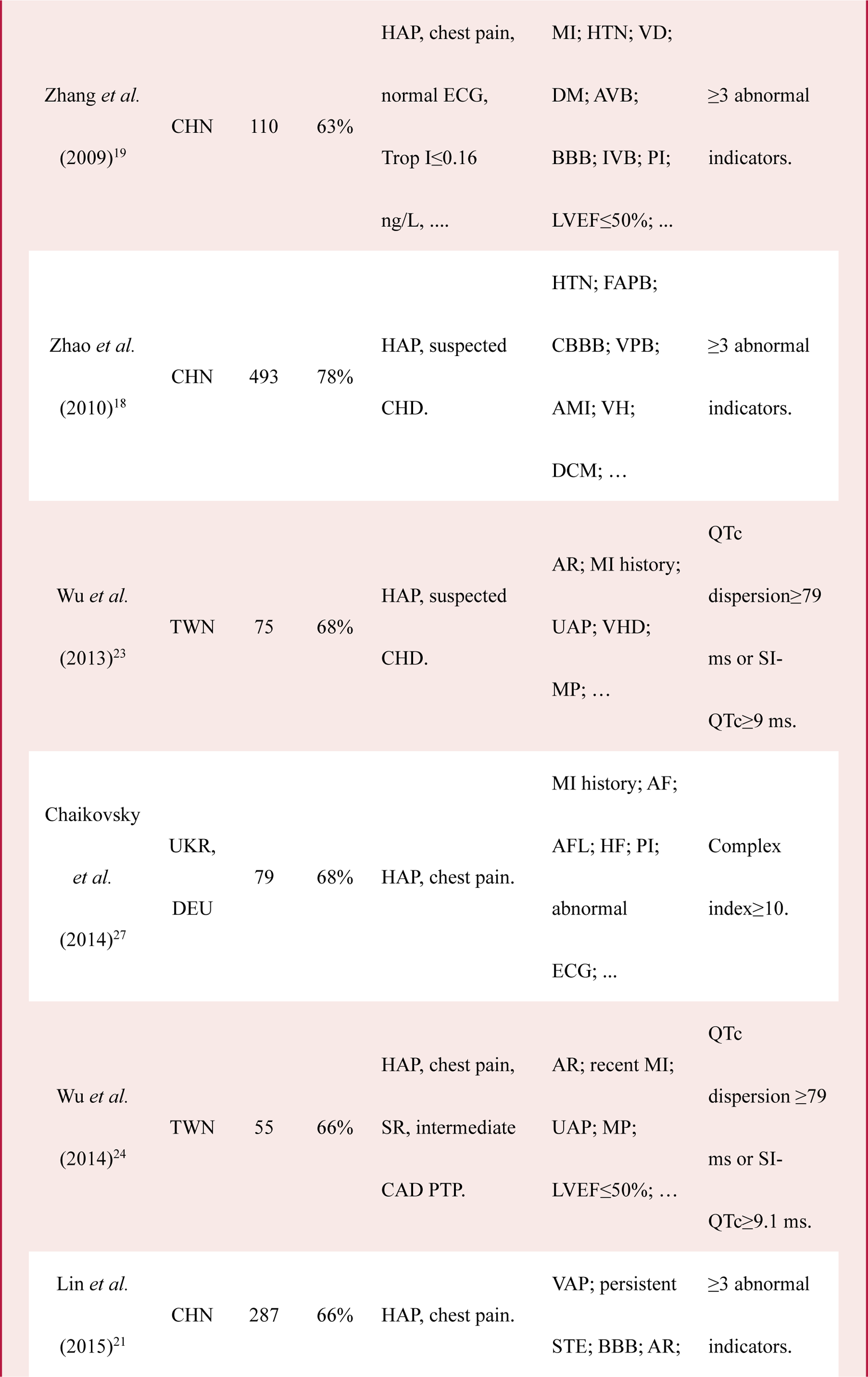
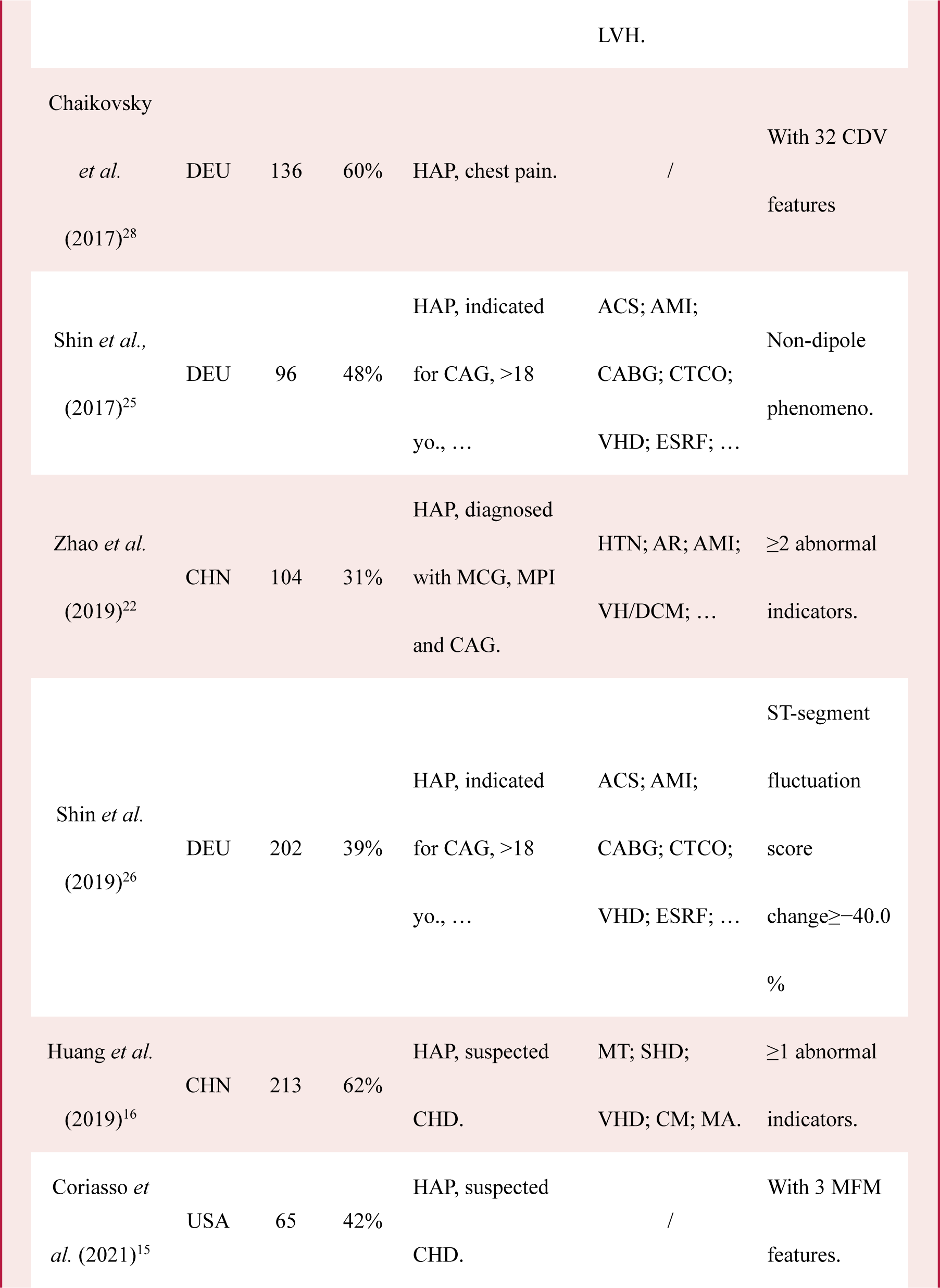
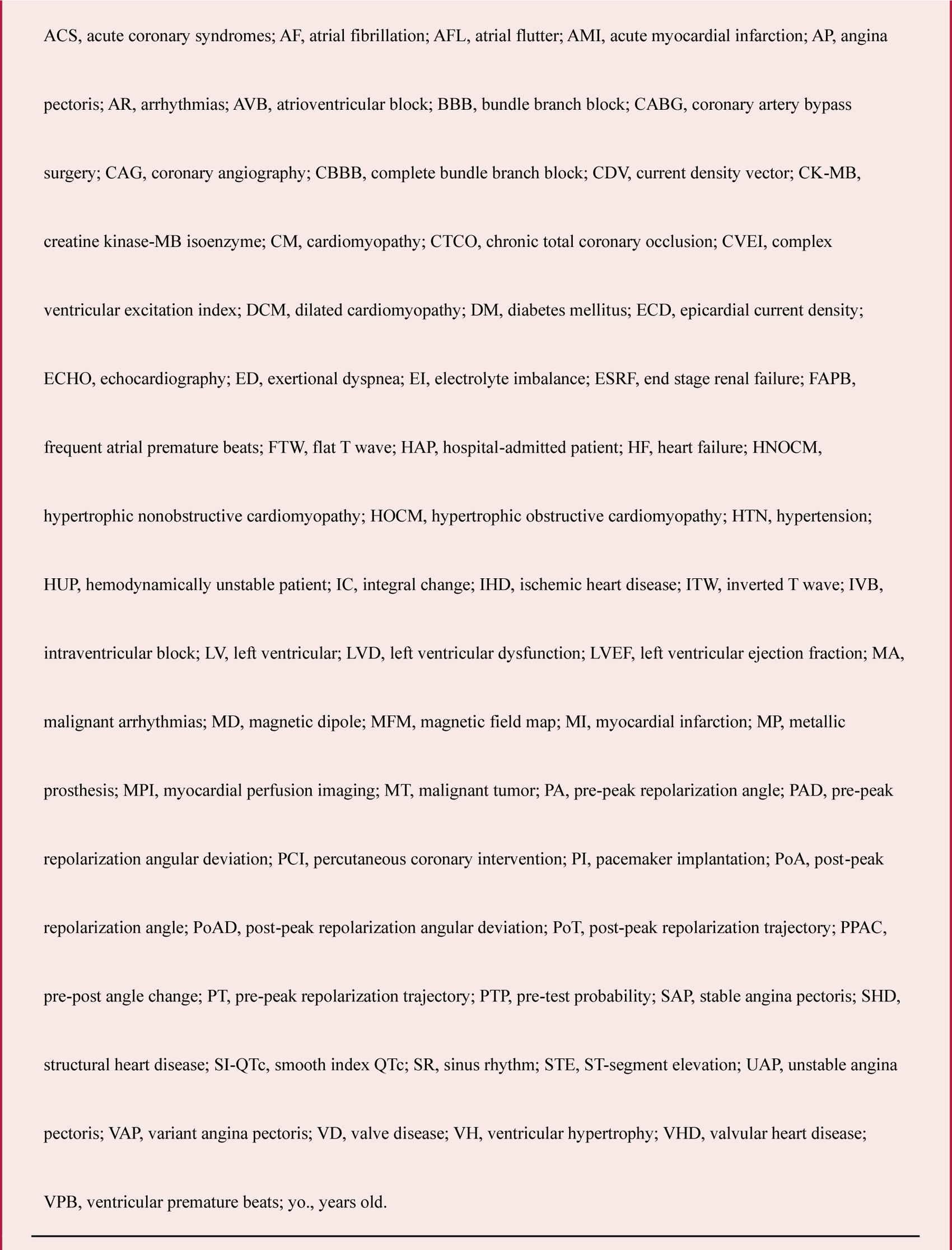
Characteristics of included studies.

The result of quality evaluation is shown in table 2 (details in supplementary materials). Due to the lack of recognized indices and the corresponding reference value, many studies choose to draw SROC curve to decide the best cut-off value, leading to high level of risk of bias in the index test. The results of each study and their combination are shown in the forest plot (figure 2). The pooled sensitivity of MCG for diagnosing CHD is 0.86 (95% CI: 0.80-0.91), and the pooled specificity is 0.79 (95% CI: 0.71-0.86). The area under SROC curve is 0.90 (95% CI: 0.87-0.92) (figure 3). The sensitivity analysis indicates the absence of significant publication bias (figure S1).

**Figure 2.**
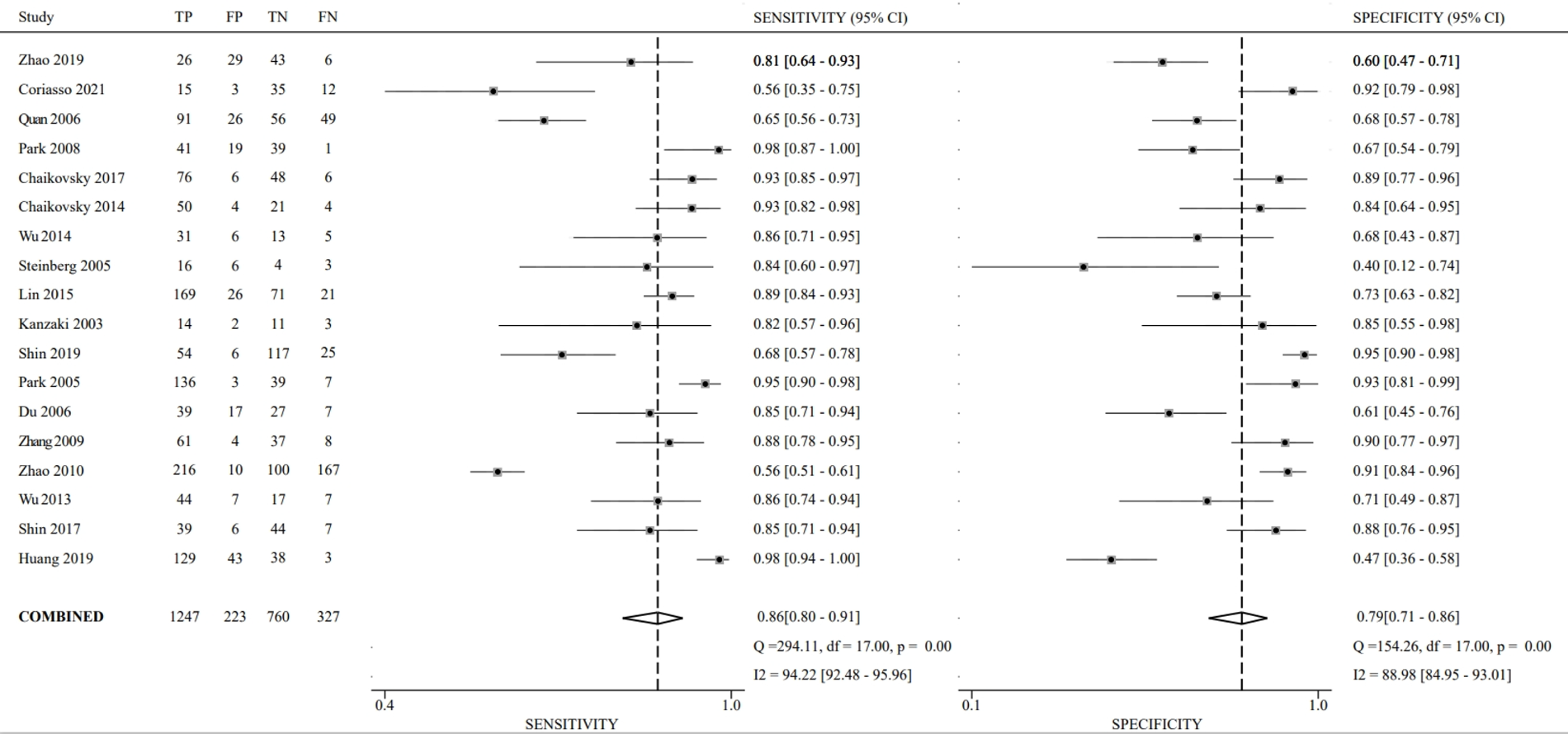
The forest plot showing the results of individual studies and their combination. The left subfigure calculates the sensitivity, and the right subfigure calculates the false positive rate. Each row in the subfigure represents a study, with the study name and the numbers of samples in diagnostic 2×2 table. The confidence interval and prediction interval for the combined index are shown at the bottom of each subfigure. The combined sensitivity of MCG for diagnosing CHD is 0.86 (0.80 – 0.91), and the combined specificity is 0.79 (0.71 – 0.86). The value of Cochran’s *Q* for sensitivity and specificity are 294.11 and 154.26, respectively. The value of *I*^2^ for sensitivity and specificity are 94.22% (92.48 – 95.96) and 88.98% (84.95 – 93.01), respectively.

**Figure 3.**
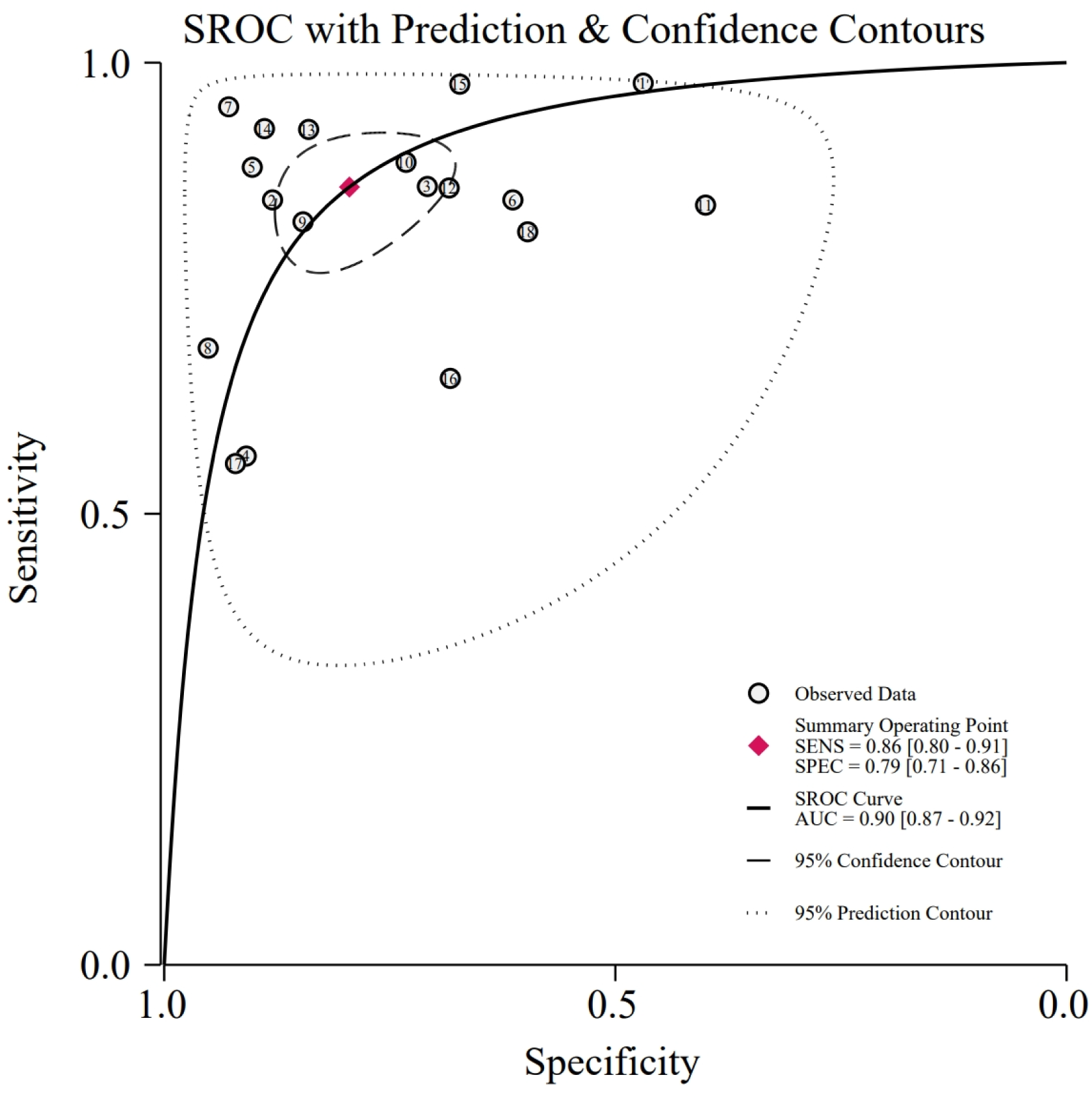
The combined results of all studies. Each red circle represents an included study, and the size of it indicates the sample size. The black line is the SROC curve, and the red star represents the summary point obtained by combination, corresponding to a sensitivity of 0.86 and a specificity of 0.79. The blue and gray dashed lines represent the confidence interval and prediction interval of the total characteristics.

**Table 2.**
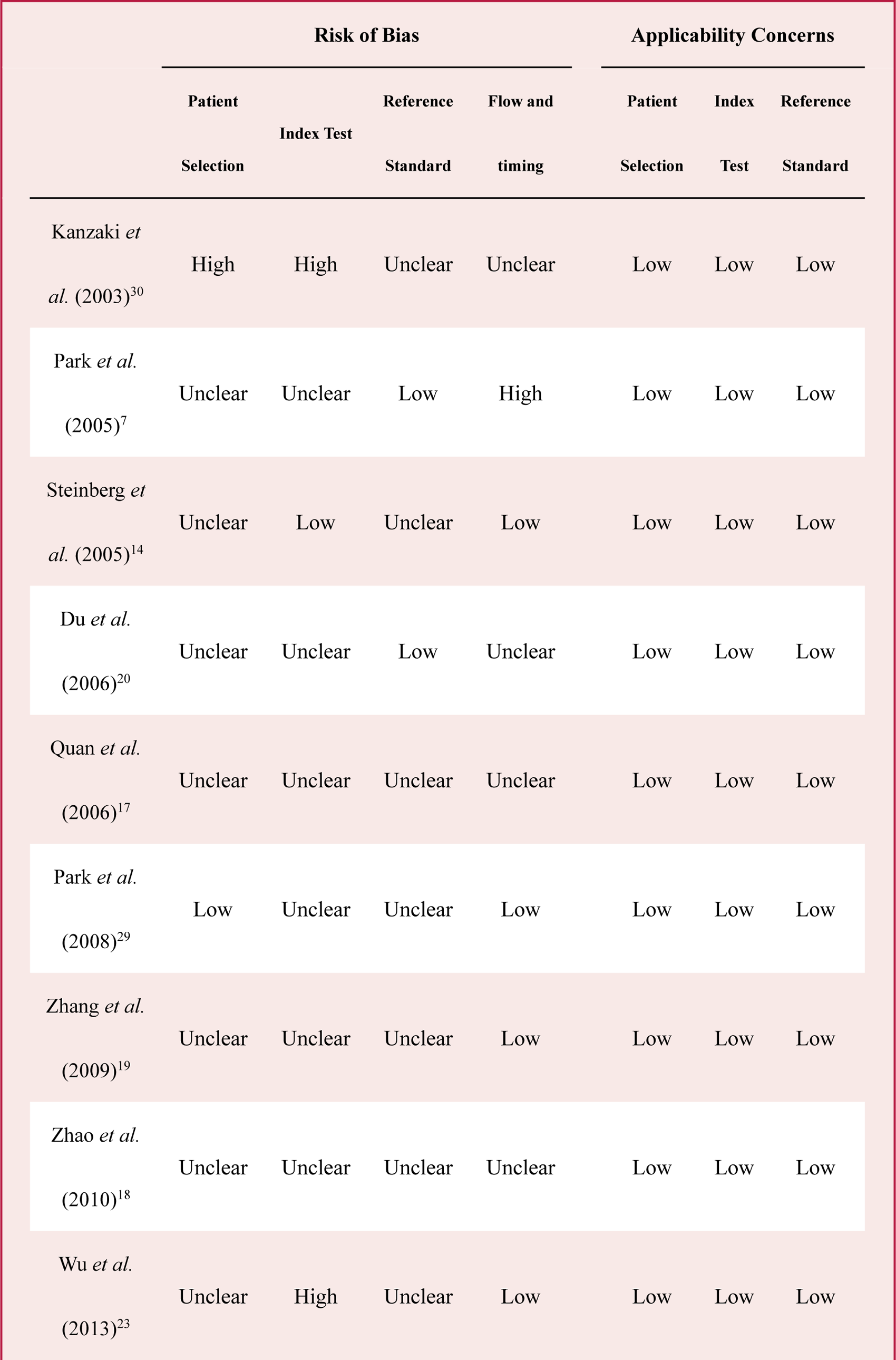
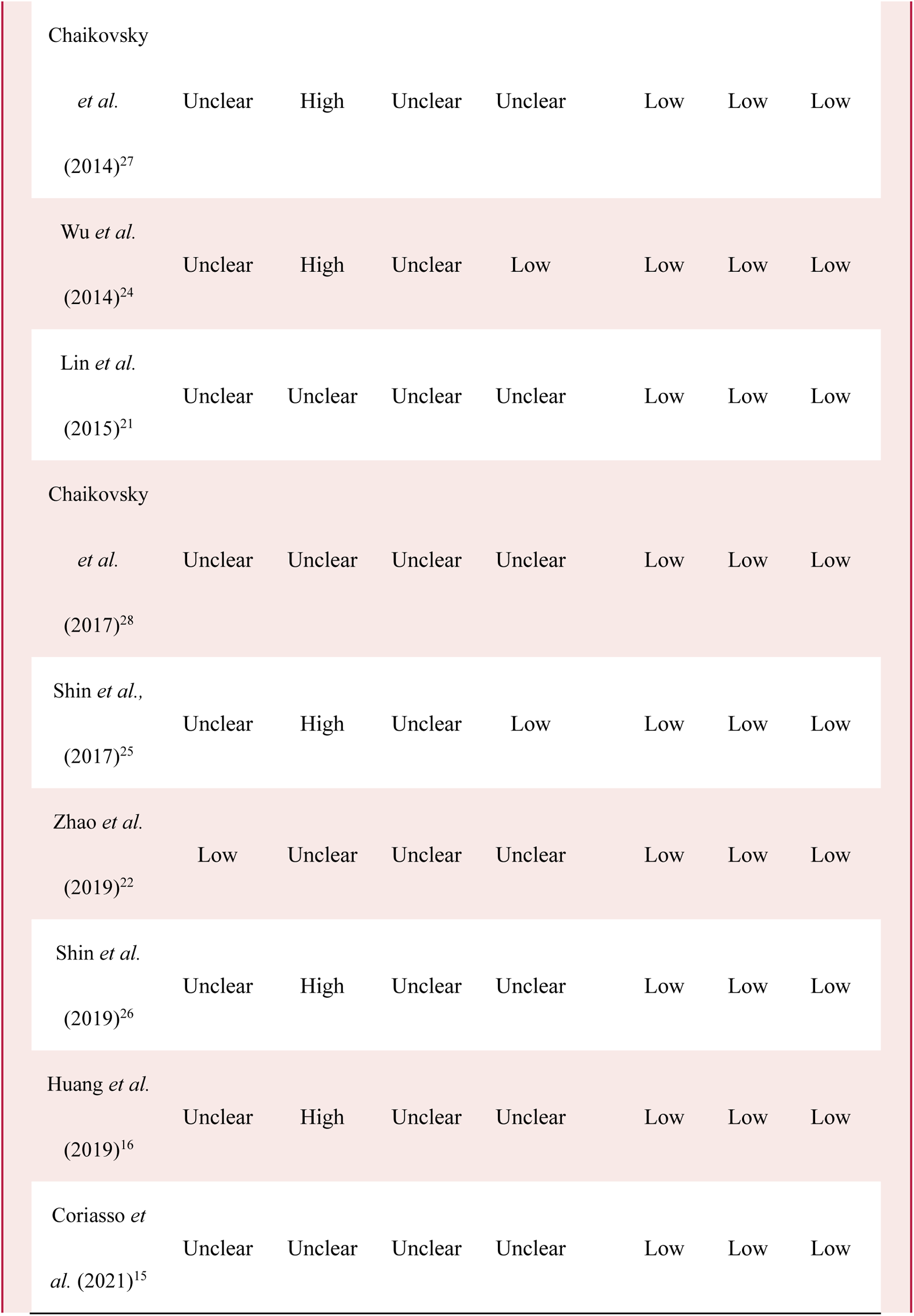
Quality evaluation. The names of each study are labeled on the left side. It encompasses the assessment of risk of bias and applicability across seven dimensions, which are evaluated on three levels: high, low, and unclear.

There is significant statistical heterogeneity between studies (*P*<0·01). The value of Cochran’s *Q* for sensitivity and specificity are 294.11 and 154.26, respectively. The *I*^2^ for sensitivity and specificity are 94.22% (95% CI: 92.48-95.96)and 88.98% (95% CI: 84.95-93.01), respectively. In the meta-regression and subgroup analysis, the value of sensitivity for mild CHD and severe CHD are 0.89 and 0.86, respectively, showing significant difference within subgroups (*P*=0.01). Other results of the analysis (table 3) are not significant due to the limited sample size.

**Table 3.** The results of meta-regression and subgroup analysis. The sensitivity for diagnosing early CHD and severe CHD performs significant difference (*P*<0 · 05). Ref-std., reference standard.

|  |  | Number of |  | Sensitivity |  | Specificity |  |
| --- | --- | --- | --- | --- | --- | --- | --- |
|  |  | Studies | Samples | Value<br>(95% CI) | p-value | Value<br>(95% CI) | p-value |
| Rest<br>ECG | Silent | 4 | 441 | 0.84<br>(0.71-0.97) | 0.09 | 0.82<br>(0.68-0.97) | 0.43 |
|  | Ensemble | 14 | 2130 | 0.87<br>(0.81-0.93) |  | 0.79<br>(0.70-0.87) |  |
| Stress<br>test | Yes | 4 | 428 | 0.86<br>(0.74-0.99) | 0.21 | 0.87<br>(0.76-0.97) | 0.71 |
|  | No | 14 | 2143 | 0.86<br>(0.80-0.92) |  | 0.77<br>(0.68-0.85) |  |
| Ref-std. | Mild | 7 | 1270 | 0.89<br>(0.83-0.96) | 0.01 | 0.78<br>(0.67-0.90) | 0.13 |
|  | Severe | 10 | 1236 | 0.86<br>(0.78-0.93) |  | 0.78<br>(0.68-0.88) |  |

## Conclusions

This article presents a systematic review and meta-analysis aimed at evaluating the performance of MCG for diagnosing CHD. The study identified 18 eligible studies, which included 2,571 subjects from 6 countries and regions. The pooled sensitivity and specificity values for MCG are 86% (95% CI: 80-91) and 79% (95% CI: 71-86), respectively, indicating that MCG is of suitable for diagnosing CHD, especially in large-scale screening. The area under the SROC curve is 0.90, implying high diagnostic value of MCG.

This article provides a comprehensive evaluation of the diagnostic performance of MCG for diagnosing CHD using high-quality clinical trials. In contrast to the study of Rajender Agarwal *et al.*^31^ which reported analogous values for pooled sensitivity and specificity, our research employs a more comprehensive information retrieval process and conducts subgroup analysis. These analysis aim to provide deeper insights into the characteristics of MCG. According to our findings, MCG has a higher sensitivity for early CHD, which increases its benefits in CHD screening. Other analyses don’t reveal any significant variations. We blame the small sample sizes in each subgroup for the issue. For instance, even though the silent group only consisted of 441 samples from 4 studies in the analysis of the patient’s resting ECG status, the p-value of 0.09 is very close to the significant level and suggests potential findings for further investigation. Future investigation of additional factors that might have an impact on MCG performance will require more research.

More opportunities for conducting large population clinical trials across the globe are made possible by the development of OPM-based MCG. However, given the lack of standardization in MCG procedures, it is crucial to optimize the design of future trials based on previous studies. The quality assessment shows unsatisfactory results concerning the risk of bias, mainly due to the unclear procedure followed in the trials. Firstly, patient selection should encompass suspected patients with varying severity levels, especially those who are challenging to diagnose accurately. Secondly, blind designs should be implemented for both CAG and MCG to minimize potential bias. Besides, to ensure reasonable flow and timing, an appropriate interval between MCG and CAG should be established. Considering the invasive nature of CAG and its potential impact on cardiac condition, MCG, as a noninvasive and quick examination, should be conducted within a 24-hour window before CAG to ensure comparable states of patients between the two tests. Additionally, in light of the absence of standardized diagnostic indices and their reference values for MCG, the existence of receiver operating characteristic (ROC) curves becomes essential to obtain sufficient values of thresholds. Large-scale global trials can be conducted based on the thresholds obtained during sufficient small-scale pre-experiment, thereby avoiding the bias introduced by the post-specified threshold.

## Panel: Research in context

### Evidence before this study

The rising global burden of coronary heart disease, with over 20 million cases in the United States in 3 years and nearly2.2 billion Canadian dollars annual hospitalization costs in Canada, underscores the urgent need for effective diagnostic methods. However, the lack of shielding methods with both high accuracy and low threshold poses a challenge. Magnetocardiography is a promising diagnostic tool that aligns with these requirements, and the recent development of unshielded OPM-based MCG devices further enhances its applicability. While many clinical trials have been conducted to obtain an accurate evaluation of MCG’s diagnostic performance, their limited sample size impacts the accuracy of the results. To solve this problem, a comprehensive evaluation through systematic review and meta-analysis is crucial to gain deeper insights into MCG’s diagnostic efficacy and drive evidence-based conclusions for its clinical application.

### Added value of this study

Our systematic review and meta-analysis show that MCG is of high diagnostic value for CHD, with a value of 86% (95% CI: 80-91) and 79% (95% CI: 71-86) for pooled sensitivity and specificity, respectively. Compared with ECG, MCG significantly enhances diagnostic accuracy while maintaining comparable specificity. Furthermore, the subgroup analysis reveals that MCG exhibits higher sensitivity for early CHD, making it particularly suitable for large-scale screening.

### Implications of all the available evidence

As MCG continues to evolve, an increase in relevant clinical trials is anticipated. Future trials should adopt a standardized design, encompass diverse patient selection, incorporate blind designs, and establish reasonable MCG-CAG intervals. Utilizing ROC curves can aid in determining diagnostic indices’ reference values for large-scale application. Additionally, integrating carbon electrodes into the MCG system may offer valuable insights for its clinical use across diverse demographics.

## Data Availability

Data sharing is not available for this article, as no data sets were generated or analyzed during the current study period.

---

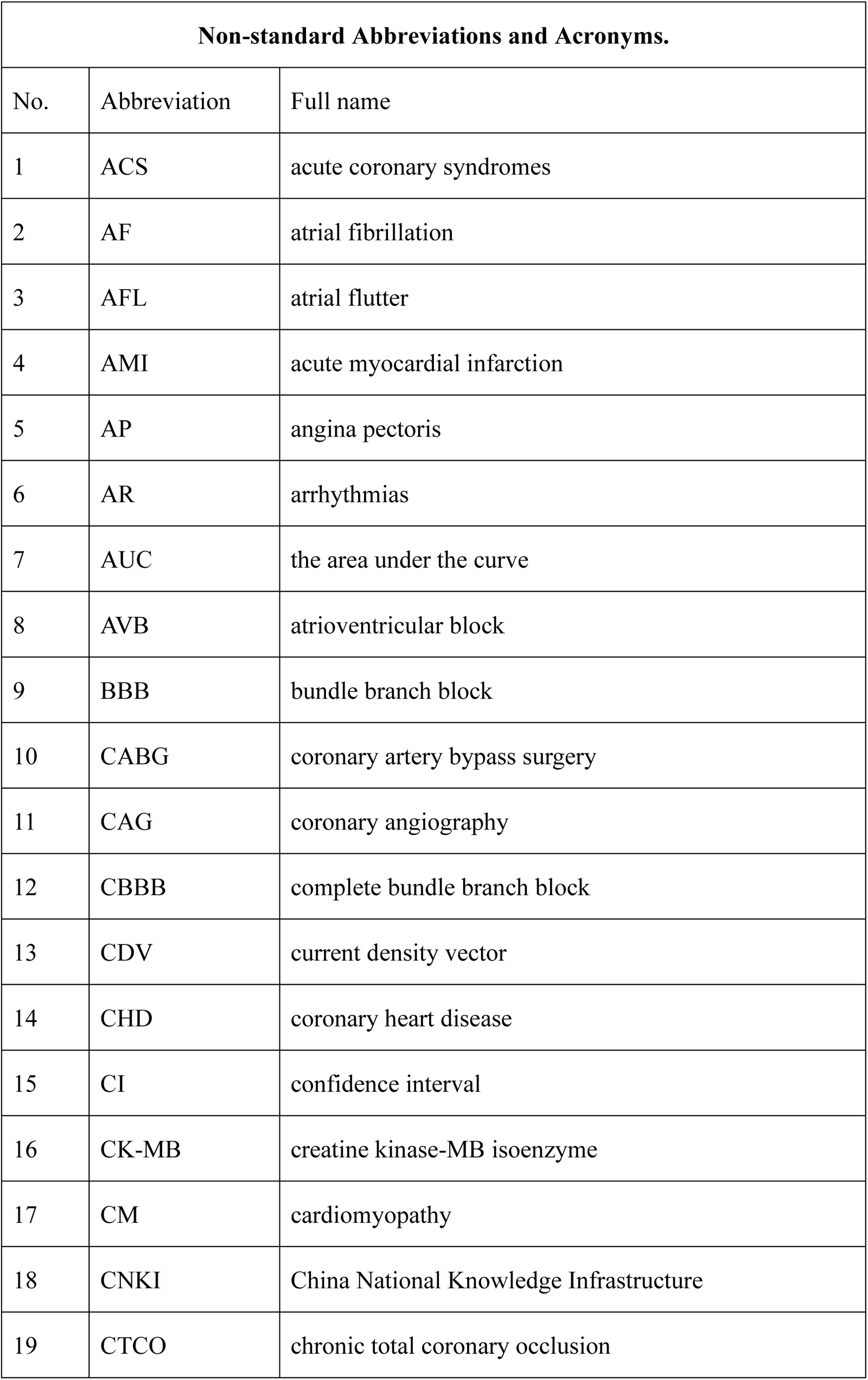
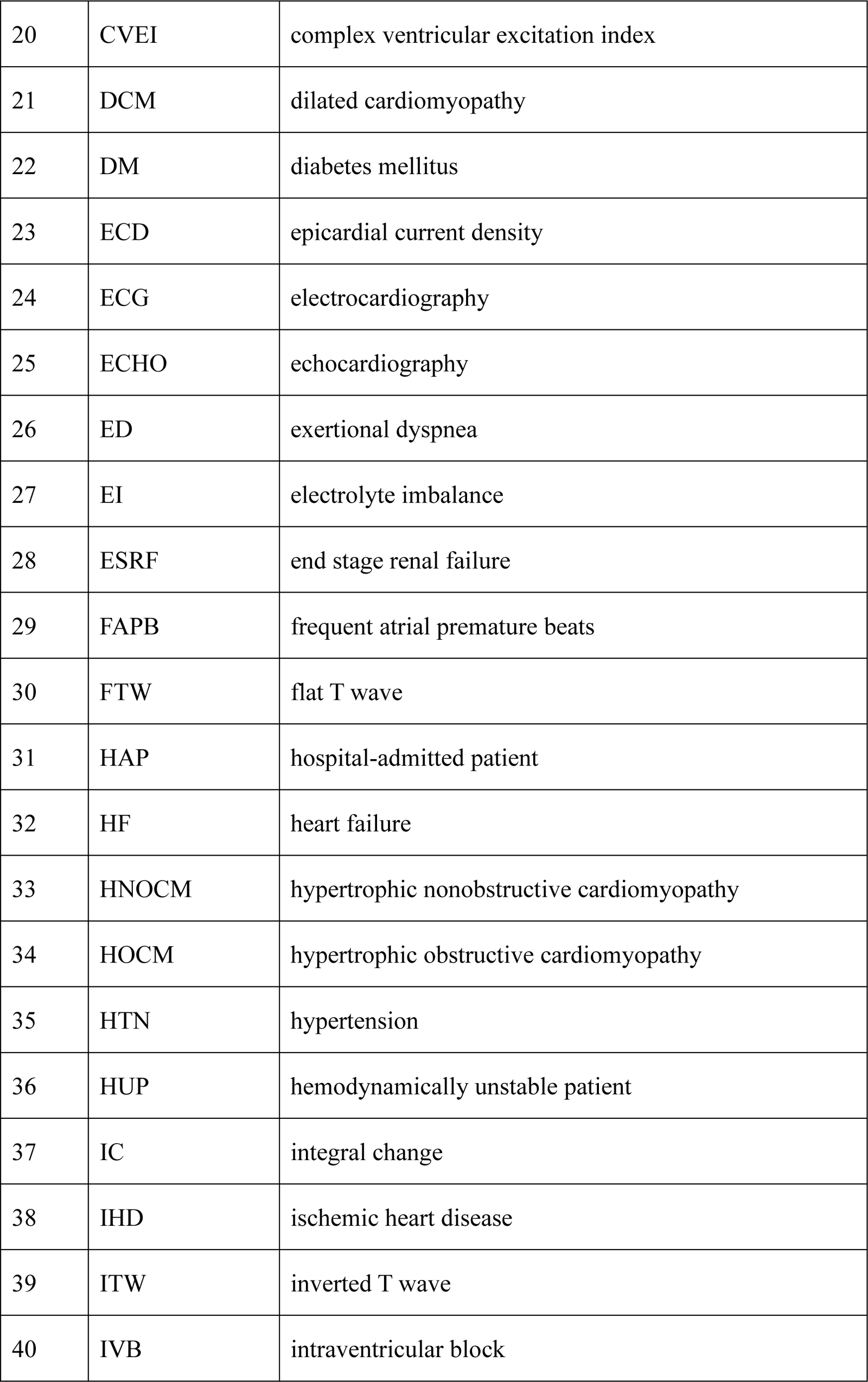
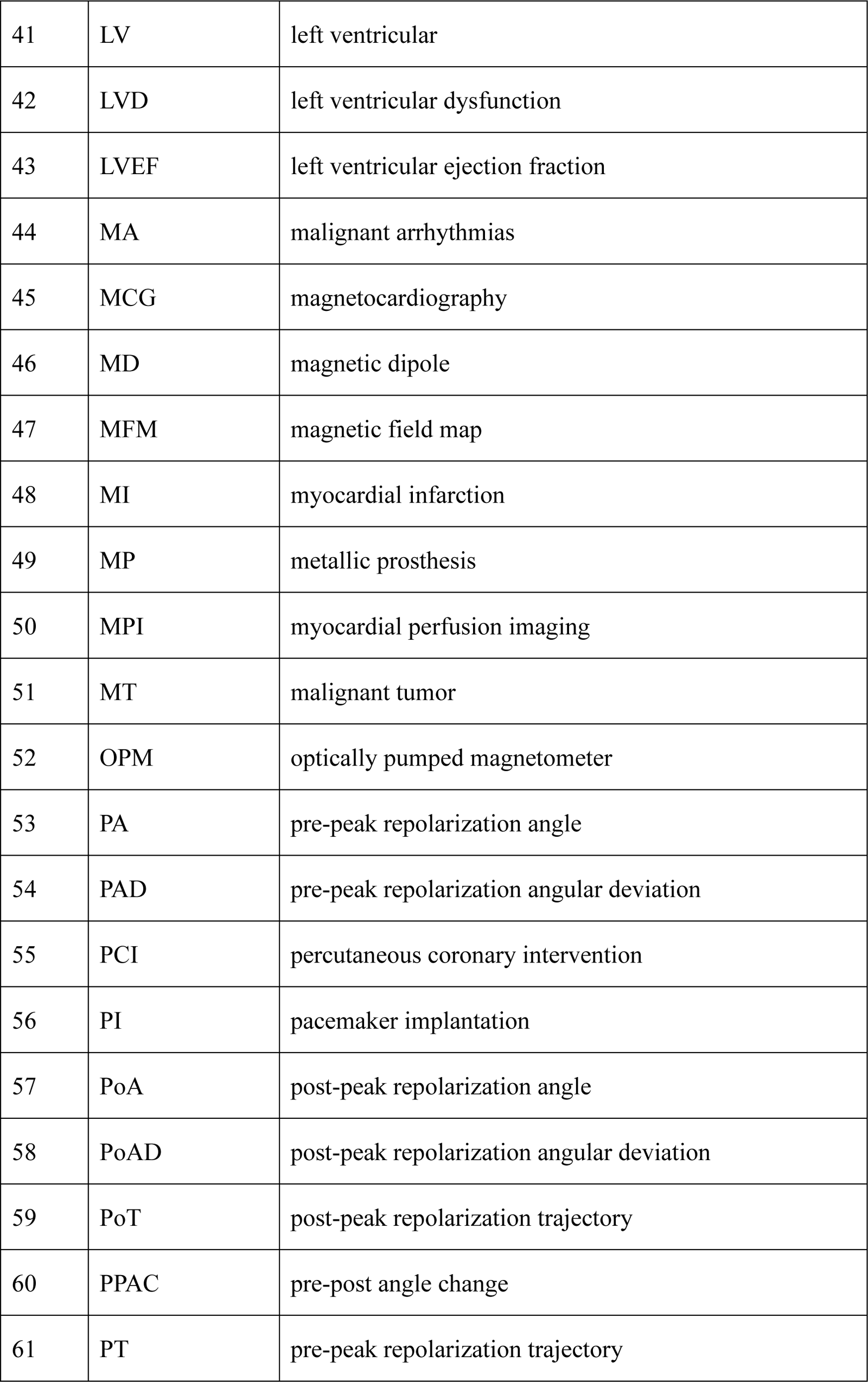
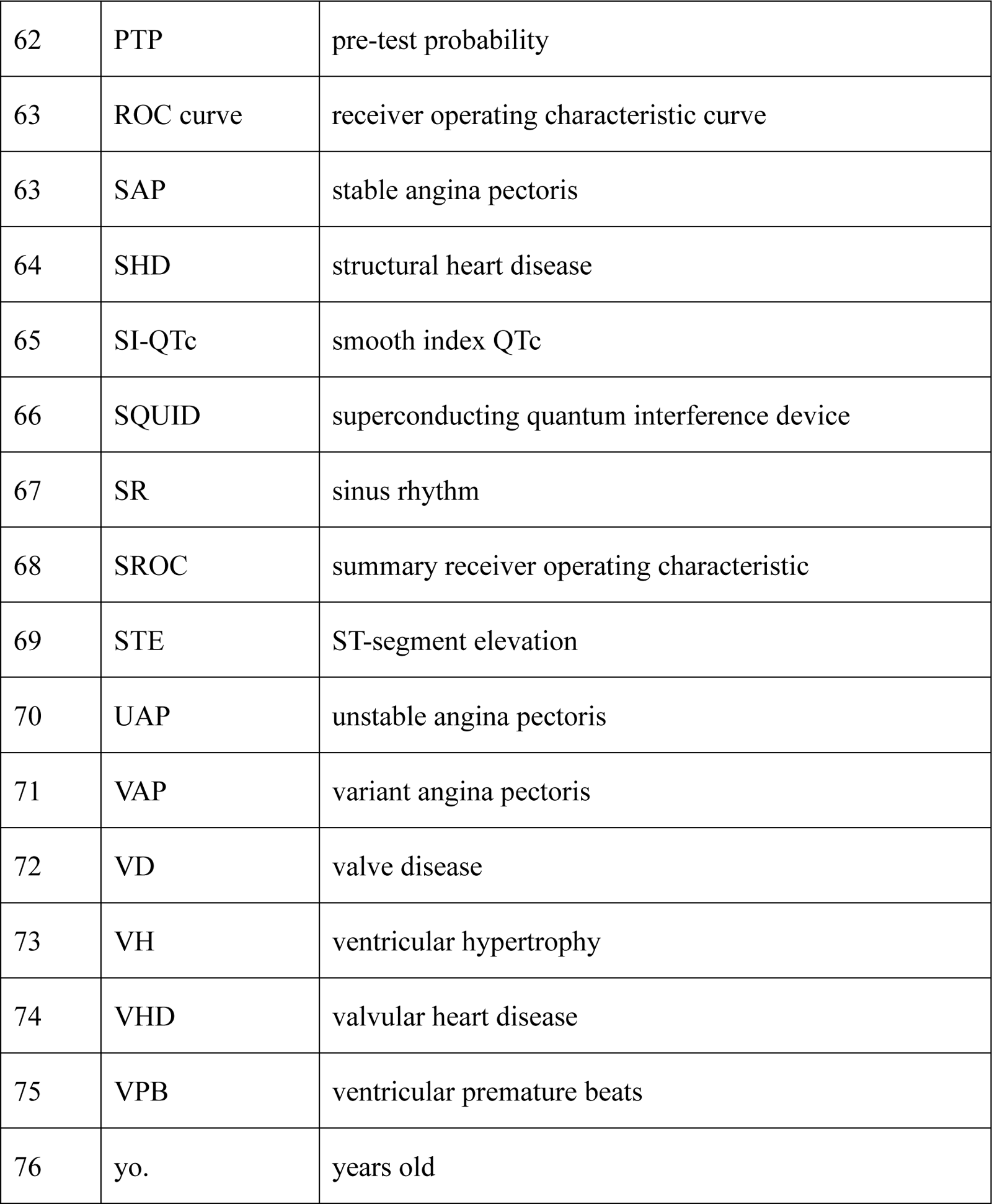

## Contributors

MY contributed to conceptualization, data accessed and verified, data analysis, data interpretation, writing—original draft, and writing—review and editing. CS contributed to conceptualization, literature search, data accessed and verified, writing—original draft, and writing—review and editing. BZ, BW, JX, MX, JZ, and TW contributed to data interpretation, and writing—review and editing. WX contributed to data interpretation, supervision, and writing—review and editing, and decision to submit. HG contributed to project administration, supervision, and writing—review and editing, and decision to submit.

## Data sharing

Extracted data and the Stata code in this study are fully reported in the supplementary material (pp. 14-16). Additional information will be available from the corresponding author upon reasonable request.

## Acknowledgements

We thank F. Li from Guanghua School of Management, Peking University for the manipulation in data analysis.

## Sources of Funding

National Natural Science Foundation of China, National Key Research and Development Program of China, Basic scientific research project of Department of Education of Liaoning Province (LJKMZ20221186), Shenyang Public Health R&D Project (22-321-33-14), 345 Talent Project .

## Disclosures

We declare no competing interests.

## Notes

### Competing Interest Statement

The authors have declared no competing interest.

### Funding Statement

Basic Scientific Research Project of Liaoning Provincial Department of Education (LJKMZ20221186), Shenyang Public Health Research and Development Project (22-321-33-14), 345 Talent Project.

