## Supplementary materials for "Magnetocardiography for the diagnosis of coronary artery disease: a systematic review and meta-analysis"

### Section 1 List of acronyms and abbreviations

| Table S1 List of acronyms and abbreviations | | |
|---|---|---|
| No. | Abbreviation | Full name |
| 1 | ACS | acute coronary syndromes |
| 2 | AF | atrial fibrillation |
| 3 | AFL | atrial flutter |
| 4 | AMI | acute myocardial infarction |
| 5 | AP | angina pectoris |
| 6 | AR | arrhythmias |
| 7 | AUC | the area under the curve |
| 8 | AVB | atrioventricular block |
| 9 | BBB | bundle branch block |
| 10 | CABG | coronary artery bypass surgery |
| 11 | CAG | coronary angiography |
| 12 | CBBB | complete bundle branch block |
| 13 | CDV | current density vector |
| 14 | CHD | coronary heart disease |
| 15 | CI | confidence interval |
| 16 | CK-MB | creatine kinase-MB isoenzyme |
| 17 | CM | cardiomyopathy |
| 18 | CNKI | China National Knowledge Infrastructure |
| 19 | CTCO | chronic total coronary occlusion |
| 20 | CVEI | complex ventricular excitation index |
| 21 | DCM | dilated cardiomyopathy |
| 22 | DM | diabetes mellitus |
| 23 | ECD | epicardial current density |
| 24 | ECG | electrocardiography |
| 25 | ECHO | echocardiography |
| 26 | ED | exertional dyspnea |
| 27 | EI | electrolyte imbalance |
| 28 | ESRF | end stage renal failure |
| 29 | FAPB | frequent atrial premature beats |
| 30 | FTW | flat T wave |
| 31 | HAP | hospital-admitted patient |
| 32 | HF | heart failure |
| 33 | HNOCM | hypertrophic nonobstructive cardiomyopathy |
| 34 | HOCM | hypertrophic obstructive cardiomyopathy |
| 35 | HTN | hypertension |
| 36 | HUP | hemodynamically unstable patient |
| 37 | IC | integral change |
| 38 | IHD | ischemic heart disease |
| 39 | ITW | inverted T wave |
| 40 | IVB | intraventricular block |
| 41 | LV | left ventricular |
| 42 | LVD | left ventricular dysfunction |

| | | |
|---|---|---|
| 43 | LVEF | left ventricular ejection fraction |
| 44 | MA | malignant arrhythmias |
| 45 | MCG | magnetocardiography |
| 46 | MD | magnetic dipole |
| 47 | MFM | magnetic field map |
| 48 | MI | myocardial infarction |
| 49 | MP | metallic prosthesis |
| 50 | MPI | myocardial perfusion imaging |
| 51 | MT | malignant tumor |
| 52 | OPM | optically pumped magnetometer |
| 53 | PA | pre-peak repolarization angle |
| 54 | PAD | pre-peak repolarization angular deviation |
| 55 | PCI | percutaneous coronary intervention |
| 56 | PI | pacemaker implantation |
| 57 | PoA | post-peak repolarization angle |
| 58 | PoAD | post-peak repolarization angular deviation |
| 59 | PoT | post-peak repolarization trajectory |
| 60 | PPAC | pre-post angle change |
| 61 | PT | pre-peak repolarization trajectory |
| 62 | PTP | pre-test probability |
| 63 | ROC curve | receiver operating characteristic curve |
| 63 | SAP | stable angina pectoris |
| 64 | SHD | structural heart disease |
| 65 | SI-QTc | smooth index QTc |
| 66 | SQUID | superconducting quantum interference device |
| 67 | SR | sinus rhythm |
| 68 | SROC | summary receiver operating characteristic |
| 69 | STE | ST-segment elevation |
| 70 | UAP | unstable angina pectoris |
| 71 | VAP | variant angina pectoris |
| 72 | VD | valve disease |
| 73 | VH | ventricular hypertrophy |
| 74 | VHD | valvular heart disease |
| 75 | VPB | ventricular premature beats |
| 76 | yo. | years old |

**Section 2 PRISMA checklist**

This research complies with the PRISMA 2020 guidelines. Details are reported at the list below.

| Section and Topic | Item # | Checklist item | Location where item is reported |
|---|---|---|---|
| **TITLE** | | | |
| Title | 1 | Identify the report as a systematic review. | Paper title |
| **ABSTRACT** | | | |
| Abstract | 2 | See the PRISMA 2020 for Abstracts checklist. | |
| **INTRODUCTION** | | | |
| Rationale | 3 | Describe the rationale for the review in the context of existing knowledge. | Main text introduction, paragraph 1-2. |
| Objectives | 4 | Provide an explicit statement of the objective(s) or question(s) the review addresses. | Main text introduction, paragraph 2. |
| **METHODS** | | | |
| Eligibility criteria | 5 | Specify the inclusion and exclusion criteria for the review and how studies were grouped for the syntheses. | Main text methods, "Search strategy and selection criteria" section. |
| Information sources | 6 | Specify all databases, registers, websites, organisations, reference lists and other sources searched or consulted to identify studies. Specify the date when each source was last searched or consulted. | Main text methods, "Search strategy and selection criteria" section. |
| Search strategy | 7 | Present the full search strategies for all databases, registers and websites, including any filters and limits used. | Supplementary material, "Search Strategy" section. |
| Selection process | 8 | Specify the methods used to decide whether a study met the inclusion criteria of the review, including how many reviewers screened each record and each report retrieved, whether they worked independently, and if applicable, details of automation tools used in the process. | Main text methods, "Data extraction" section. |
| Data collection process | 9 | Specify the methods used to collect data from reports, including how many reviewers collected data from each report, whether they worked independently, any processes for obtaining or confirming data from study investigators, and if applicable, details of automation tools used in the process. | Main text methods, "Data extraction" section. |
| Data items | 10a | List and define all outcomes for which data were sought. Specify whether all results that were compatible with each outcome domain in each study were sought (e.g. for all measures, time points, analyses), and if not, the methods used to decide | Main text methods, "Data extraction" section.<br>Supplementary material "Data extraction" section. |

| Section and Topic | Item # | Checklist item | Location where item is reported |
|---|---|---|---|
| | | which results to collect. | |
| | 10b | List and define all other variables for which data were sought (e.g. participant and intervention characteristics, funding sources). Describe any assumptions made about any missing or unclear information. | Main text methods, “Data extraction” section. |
| Study risk of bias assessment | 11 | Specify the methods used to assess risk of bias in the included studies, including details of the tool(s) used, how many reviewers assessed each study and whether they worked independently, and if applicable, details of automation tools used in the process. | Main text, “Quality evaluation” section.<br>Supplementary material, “Quality evaluation” section. |
| Effect measures | 12 | Specify for each outcome the effect measure(s) (e.g. risk ratio, mean difference) used in the synthesis or presentation of results. | Main text methods, “Data analysis” section. |
| Synthesis methods | 13a | Describe the processes used to decide which studies were eligible for each synthesis (e.g. tabulating the study intervention characteristics and comparing against the planned groups for each synthesis (item #5)). | Main text methods, “Search strategy and selection criteria” section. |
| | 13b | Describe any methods required to prepare the data for presentation or synthesis, such as handling of missing summary statistics, or data conversions. | Main text methods, “Data extraction”.<br>Main text methods, “Data analysis”. |
| | 13c | Describe any methods used to tabulate or visually display results of individual studies and syntheses. | Main text methods, “Data analysis”. |
| | 13d | Describe any methods used to synthesize results and provide a rationale for the choice(s). If meta-analysis was performed, describe the model(s), method(s) to identify the presence and extent of statistical heterogeneity, and software package(s) used. | Main text methods, “Data analysis”. |
| | 13e | Describe any methods used to explore possible causes of heterogeneity among study results (e.g. subgroup analysis, meta-regression). | Main text methods, “Data analysis”. |
| | 13f | Describe any sensitivity analyses conducted to assess robustness of the synthesized results. | Supplementary materials, “sensitivity analysis” section. |
| Reporting bias assessment | 14 | Describe any methods used to assess risk of bias due to missing results in a synthesis (arising from reporting biases). | / |
| Certainty assessment | 15 | Describe any methods used to assess certainty (or confidence) in the body of evidence for an outcome. | / |

| Section and Topic | Item # | Checklist item | Location where item is reported |
|---|---|---|---|
| **RESULTS** | | | |
| Study selection | 16a | Describe the results of the search and selection process, from the number of records identified in the search to the number of studies included in the review, ideally using a flow diagram. | Main text results, paragraph 1-2. |
| | 16b | Cite studies that might appear to meet the inclusion criteria, but which were excluded, and explain why they were excluded. | Supplementary materials, "Selection criteria" section. |
| Study characteristics | 17 | Cite each included study and present its characteristics. | Main text, table 1. |
| Risk of bias in studies | 18 | Present assessments of risk of bias for each included study. | Main text results, paragraph 3. |
| Results of individual studies | 19 | For all outcomes, present, for each study: (a) summary statistics for each group (where appropriate) and (b) an effect estimate and its precision (e.g. confidence/credible interval), ideally using structured tables or plots. | Main text, figure 2. |
| Results of syntheses | 20a | For each synthesis, briefly summarise the characteristics and risk of bias among contributing studies. | Main text results, paragraph 3. |
| | 20b | Present results of all statistical syntheses conducted. If meta-analysis was done, present for each the summary estimate and its precision (e.g. confidence/credible interval) and measures of statistical heterogeneity. If comparing groups, describe the direction of the effect. | Main texts results, paragraph 3. |
| | 20c | Present results of all investigations of possible causes of heterogeneity among study results. | Main texts results, paragraph 4. |
| | 20d | Present results of all sensitivity analyses conducted to assess the robustness of the synthesized results. | Supplementary materials, "Sensitivity analysis" section. |
| Reporting biases | 21 | Present assessments of risk of bias due to missing results (arising from reporting biases) for each synthesis assessed. | / |
| Certainty of evidence | 22 | Present assessments of certainty (or confidence) in the body of evidence for each outcome assessed. | For all mean estimates: 95% confidence intervals (CIs). |
| **DISCUSSION** | | | |
| Discussion | 23a | Provide a general interpretation of the results in the context of other evidence. | Main texts discussion, paragraph 1. |
| | 23b | Discuss any limitations of the evidence included in the review. | Main texts discussion, paragraph 2. |

| Section and Topic | Item # | Checklist item | Location where item is reported |
|---|---|---|---|
| | 23c | Discuss any limitations of the review processes used. | Main texts discussion, paragraph 2. |
| | 23d | Discuss implications of the results for practice, policy, and future research. | Main texts discussion, paragraph 4-5. |
| **OTHER INFORMATION** | | | |
| Registration and protocol | 24a | Provide registration information for the review, including register name and registration number, or state that the review was not registered. | Abstract methods. |
| | 24b | Indicate where the review protocol can be accessed, or state that a protocol was not prepared. | Main texts, data sharing. |
| | 24c | Describe and explain any amendments to information provided at registration or in the protocol. | / |
| Support | 25 | Describe sources of financial or non-financial support for the review, and the role of the funders or sponsors in the review. | Main texts methods, Role of the funding source. |
| Competing interests | 26 | Declare any competing interests of review authors. | Main texts, Declaration of interests. |
| Availability of data, code and other materials | 27 | Report which of the following are publicly available and where they can be found: template data collection forms; data extracted from included studies; data used for all analyses; analytic code; any other materials used in the review. | Main texts, data sharing. |

### Section 3 Search strategy

PubMed

| Table S2 Search strategy for PubMed | |
|---|---|
| No. | Query |
| #5 | #1 AND #2 AND #3 AND #4 |
| #4 | ((predictive[Title/Abstract]) AND (value[Title/Abstract])) OR ((sensitiv*[Title/Abstract]) OR (sensitivity and specificity[MeSH Terms])) |
| #3 | angiogra*[Title/Abstract] |
| #2 | ((((((((((((((Coronary Artery Disease[Title/Abstract]) OR (Arteriosclerosis, Coronary[Title/Abstract])) OR (Coronary Atherosclerosis[Title/Abstract])) OR (Coronary Atheroscleroses[Title/Abstract])) OR (Atheroscleroses, Coronary[Title/Abstract])) OR (Atherosclerosis, Coronary[Title/Abstract])) OR (Coronary Arterioscleroses[Title/Abstract])) OR (Arterioscleroses, Coronary[Title/Abstract])) OR (Coronary Arteriosclerosis[Title/Abstract])) OR (Left Main Coronary Disease[Title/Abstract])) OR (Left Main Diseases[Title/Abstract])) OR (Left Main Disease[Title/Abstract])) OR (Left Main Coronary Artery Disease[Title/Abstract])) OR (Coronary Artery Diseases[Title/Abstract])) OR (Artery Diseases, Coronary[Title/Abstract])) OR (Artery Disease, Coronary[Title/Abstract]) |
| #1 | (Magnetocardiography[Title/Abstract]) OR (magnetocardiogra*[Title/Abstract]) |
| Number of studies: 17 | |

Embase

| Table S3 Search strategy for Embase | |
|---|---|
| No. | Query |
| #5 | #1 AND #2 AND #3 AND #4 |
| #4 | 'angiography*':ab,ti |
| #3 | 'sensitiv':ab,ti OR 'diagnostic accuracy':ab,ti OR 'diagnostic':ab,ti |
| #2 | 'magnetocardiogra*':ab,ti OR 'magnetocardiography'/exp |
| #1 | 'arteriosclerosis, coronary':ab,ti OR 'coronary atherosclerosis':ab,ti OR 'coronary atheroscleroses':ab,ti OR 'atheroscleroses, coronary':ab,ti OR 'atherosclerosis, coronary':ab,ti OR 'coronary arterioscleroses':ab,ti OR 'arterioscleroses, coronary':ab,ti OR 'coronary arteriosclerosis':ab,ti OR 'left main coronary disease':ab,ti OR 'left main diseases':ab,ti OR 'left main disease':ab,ti OR 'left main coronary artery disease':ab,ti OR 'coronary artery diseases':ab,ti OR 'artery diseases, coronary':ab,ti OR 'artery disease, coronary':ab,ti OR 'coronary artery disease'/exp |
| Number of studies: 13 | |

Web of Science

| Table S4 Search strategy for Web of Science | |
|---|---|
| No. | Query |
| 1 | TS=(Coronary Artery disease OR Artery Disease OR Coronary Left Main Diseases OR Atherosclerosis, Coronary OR Artery Diseases, Coronary OR Left Main Coronary Disease OR Atheroscleroses, Coronary OR Coronary Artery Diseases OR Coronary Arteriosclerosis OR Coronary Atheroscleroses OR Left Main Coronary Artery Disease OR Arterioscleroses, Coronary OR Coronary Atherosclerosis OR Left Main Disease OR Coronary Arterioscleroses OR Arteriosclerosis, Coronary) |
| 2 | TS=(magnetocardiography OR magnetocardiogra*) |
| 3 | TS=(sensitiv* OR sensitivity and specificity OR (predictive AND value) OR predictive value of tests OR accuracy*) |

| | |
|---|---|
| 4 | TS=angiogra* |
| 5 | #4 AND #3 AND #2 AND #1 |
| Number of studies: 26 | |

Cochrane Library

| **Table S5 Search strategy for Cochrane Library** | |
|---|---|
| No. | Query |
| #1 | MeSH descriptor: [Coronary Artery Disease] explode all trees |
| #2 | (Left Main Diseases):ti,ab,kw OR (Left Main Coronary Disease):ti,ab,kw OR (Coronary Arteriosclerosis):ti,ab,kw OR (Arterioscleroses, Coronary):ti,ab,kw OR (Coronary Arterioscleroses):ti,ab,kw (Word variations have been searched) |
| #3 | (Artery Disease, Coronary):ti,ab,kw OR (Artery Diseases, Coronary):ti,ab,kw OR (Coronary Artery Diseases):ti,ab,kw OR (Left Main Coronary Artery Disease):ti,ab,kw OR (Left Main Disease):ti,ab,kw (Word variations have been searched) |
| #4 | (Atherosclerosis, Coronary):ti,ab,kw OR (Atheroscleroses, Coronary):ti,ab,kw OR (Coronary Atheroscleroses):ti,ab,kw OR (Coronary Atherosclerosis):ti,ab,kw OR (Arteriosclerosis, Coronary):ti,ab,kw (Word variations have been searched) |
| #5 | #1 OR #2 OR #3 OR #4 |
| #6 | MeSH descriptor: [Magnetocardiography] explode all trees |
| #7 | (magnetocardiogra*):ti,ab,kw (Word variations have been searched) |
| #8 | #6 OR #7 |
| #9 | #5 AND #8 |
| Number of studies: 4 | |

ClinicalTrials.gov

| **Table S6 Search strategy for ClinicalTrials.gov** | |
|---|---|
| Intervention/treatment | Magnetocardiography OR magnetocardiogram |
| Condition or disease | Coronary artery disease |
| Number of studies: 12 (44 articles mentioned) | |

Scopus

| **Table S7 Search strategy for Scopus** | |
|---|---|
| No. | Query |
| #1 | TITLE-ABS-KEY ( magnetocardiography ) OR TITLE-ABS-KEY ( magnetocardiogra* ) |
| #2 | TITLE-ABS-KEY ( angiogra* ) |
| #3 | TITLE-ABS-KEY ( sensitiv* ) OR TITLE-ABS-KEY ( sensitivity AND specificity ) OR TITLE-ABS-KEY ( predictive AND value ) OR TITLE-ABS-KEY ( predictive AND value AND of AND tests ) OR TITLE-ABS-KEY ( accuracy* ) |
| #4 | TITLE-ABS-KEY ( artery AND disease, AND coronary ) OR TITLE-ABS-KEY ( artery AND diseases, AND coronary ) OR TITLE-ABS-KEY ( coronary AND artery AND diseases ) OR TITLE-ABS-KEY ( left AND main AND coronary AND artery AND disease ) OR TITLE-ABS-KEY ( left AND main AND disease ) OR TITLE-ABS-KEY ( left AND main AND diseases ) OR TITLE-ABS-KEY ( left AND main AND coronary AND disease ) OR TITLE-ABS-KEY ( coronary AND arteriosclerosis ) OR TITLE-ABS-KEY ( arterioscleroses, AND coronary ) OR TITLE-ABS-KEY ( coronary AND arterioscleroses ) OR TITLE-ABS-KEY ( atherosclerosis, AND coronary ) OR TITLE-ABS-KEY ( atheroscleroses, AND coronary ) OR TITLE-ABS-KEY ( coronary AND atheroscleroses ) OR TITLE-ABS-KEY ( coronary AND atherosclerosis ) OR TITLE-ABS-KEY ( arteriosclerosis, AND coronary ) OR TITLE-ABS-KEY ( coronary AND artery AND disease ) |
| #5 | #1 AND #2 AND #3 AND #4 |

| Number of studies: 33 |
| --- |

CNKI

| Table S8 Search strategy for CNKI | |
| --- | --- |
| No. | Topic words |
| 1 | 冠心病 |
| 2 | 冠状动脉 |
| 3 | 心肌缺血 |
| 4 | 缺血性心脏病 |
| 5 | 1 OR 2 OR 3 OR 4 |
| 6 | 心磁 |
| 7 | 诊断 |
| 8 | 冠状动脉造影 |
| 9 | 冠脉造影 |
| 10 | 8 OR 9 |
| 11 | 5 AND 6 AND 7 AND 10 |
| Number of studies: 10 | |

Wanfang

| Table S9 Search strategy for Wanfang | |
| --- | --- |
| # | Topic words |
| 1 | 冠心病 |
| 2 | 冠状动脉 |
| 3 | 心肌缺血 |
| 4 | 缺血性心脏病 |
| 5 | 1 OR 2 OR 3 OR 4 |
| 6 | 心磁 |
| 7 | 诊断 |
| 8 | 冠状动脉造影 |
| 9 | 冠脉造影 |
| 10 | 8 OR 9 |
| 11 | 5 AND 6 AND 7 AND 10 |
| Number of studies: 27 | |

### Section 4 Selection criteria

| Table S10 Eliminated studies during the screening | | | | |
|---|---|---|---|---|
| No. | Research | Author | Year | Reasons for elimination |
| 1 | MCG-Net: End-to-End Fine-Grained Delineation and Diagnostic Classification of Cardiac Events from Magnetocardiographs | Tao R, Zhang S, Wang Y, Mi X, Ma J, Shen C, et al. | 2022 | Different diseases; machine learning. |
| 2 | Magnetocardiography as a noninvasive diagnostic strategy for suspected coronary microvascular dysfunction | Quesada O, Pico M, Palmer C, Yildiz M, Miranda R, Malhotra R, et al. | 2022 | Immature machine learning |
| 3 | Detection of coronary artery disease in patients with chest pain: A machine learning model based on magnetocardiography parameters | Huang X, Chen P, Tang F, and Hua N | 2021 | Immature machine learning |
| 4 | Value of magnetocardiography in chronic coronary disease detection: results of multicenter trial | Chaikovsky I, Li T, Zhang W, Kazmirchyk A, Mjasnikov G, Lutay M, et al. | 2021 | Unsatisfactory subjects |
| 5 | JCS 2018 guideline on diagnosis of chronic coronary heart diseases | Yamagishi M, Tamaki N, Akasaka T, Ikeda T, Ueshima K, Uemura S, et al. | 2021 | Not a DT |
| 6 | Safety and efficacy of the European Society of Cardiology 0/1-hour algorithm for diagnosis of myocardial infarction: systematic review and meta-analysis. | Chiang CH, Chiang CH, Lee GH, et al. | 2020 | Not a DT |
| 7 | High-sensitivity-cardiac troponin for accelerated diagnosis of acute myocardial infarction: A systematic review and meta-analysis | Lee CC, Huang SS, Yeo YH, et al. | 2020 | Not a DT |
| 8 | Cardiac troponin had limited diagnostic value for acute myocardial infarction in renal insufficiency: a meta-analysis | Yang G, Yao Y, Du Y, and Huang J | 2020 | Not a DT |
| 9 | Effectiveness of magnetocardiography to identify patients in need of coronary artery revascularization: a cross-sectional study | Huang X, Hua N, Tang F, and Zhang S | 2020 | Different disease |
| 10 | A 90-second magnetocardiogram using a novel analysis system to assess for coronary artery stenosis in Emergency department observation unit chest pain patients | Pena ME, Pearson CL, Goulet MP, Kazan VM, DeRita AL, Szpunar SM, et al. | 2020 | Different disease |
| 11 | Magnetocardiography for identification of coronary ischemia in patients with chest pain and normal resting 12-lead electrocardiogram | Ramesh R, enthilnathan S, Satheesh S, Swain PP, Patel R, Ananthakrishna Pillai A, et al. | 2020 | Unsatisfactory reference standard |
| 12 | Emergency Department Observation Unit Utilization Among Older Patients With Chest Pain | Madsen T, Perkins R, Holt B, et al. | 2019 | Not a DT |
| 13 | Magnetocardiography-Based Ischemic Heart Disease Detection and Localization Using Machine Learning Methods | Tao R, Zhang S, Huang X, Tao M, Ma J, Ma S, et al. | 2019 | Immature machine learning |
| 14 | Heart Disease and Stroke Statistics-2017 Update: A Report From the American Heart Association. | Benjamin EJ, Blaha MJ, Chiuve SE, et al. | 2017 | Not a DT |
| 15 | Coronary computed tomographic angiography for patients with low-to-intermediate risk chest pain: A systematic review and meta-analysis | Chen Y, Fan Y, Yin Z, et al. | 2017 | Not a DT |
| 16 | Machine learning for prediction of all-cause mortality in patients with suspected coronary artery disease: a 5-year multicentre prospective registry analysis | Motwani M, Dey D, Berman DS, et al. | 2017 | Not a DT |
| 17 | 四通道无屏蔽心磁图仪对冠心病的诊断研究（Study on the Diagnosis of Coronary Heart Disease with Four Channel Non-shielded Magnetocardiography) | Dai Z | 2017 | Unsatisfactory reference standard. |
| 18 | Repolarization Heterogeneity of Magnetocardiography Predicts Long-Term Prognosis in Patients with Acute Myocardial Infarction | Bang WD, Kim K, Lee YH, et al. | 2016 | Not a DT; different disease |

| | | | | |
|---|---|---|---|---|
| 19 | Repolarization Heterogeneity of Magnetocardiography Predicts Long-Term Prognosis in Patients with Acute Myocardial Infarction | Her AY and Park JW | 2016 | Not a DT |
| 20 | Diagnostic outcomes of magnetocardiography in patients with coronary artery disease | Li Y, Che Z, Quan W, et al. | 2015 | Unsatisfactory subjects |
| 21 | Significance of an Indeterminate Troponin I in Patients Evaluated for Chest Pain in an Emergency Department Observation Unit | Madsen TE, Stewart M, Smyres C, Beal A, Hamilton D, Vlasic K, and Oates A. | 2015 | Not MCG |
| 22 | Validation of magnetocardiography versus fractional flow reserve for detection of coronary artery disease | Park JW, Shin ES, Ann SH, Goedde M, Park LSI, Brachmann J, et al. | 2015 | Unsatisfactory reference standard |
| 23 | Noninvasively diagnosing coronary artery disease with 61 channel MCG data | Chen T, Zhao C, Jiang S, Van Leeuwen P, and Grönemeyer D | 2014 | Unsatisfactory subjects |
| 24 | 高温超导心磁图仪在糖尿病患者冠脉介入治疗中的应用（Value of High Temperature Superconductor Magnetocardiography in Percutaneous Coronary Intervention in Patients with Type II Diabetes) | Di C, Hua N, Xie J, Tao Y, Wang B, and Tang F | 2014 | Different disease |
| 25 | Diagnostic value of magnetocardiography in coronary artery disease and cardiac arrhythmias: a review of clinical data | Kwong JS, Leithauser B, Park JW, and Yu CM | 2013 | Not a DT |
| 26 | 心磁图在心血管疾病的临床应用研究（The clinical application of magnetocardiography in cardiovascular disease) | Tao Y, Tang F, and Wang L | 2013 | Not a DT |
| 27 | 高温超导心磁图仪在冠心病中的应用初探（The value of high temperature superconductor magnetocardiography in coronary artery disease) | Di C, Hua N, Lin L, Tang F, Ma P, and Yang T | 2013 | Not a DT |
| 28 | Magnetocardiography for the diagnosis of coronary artery disease: a systematic review and meta-analysis | Agarwal R, Saini , Alyousef T, and Umscheid CA | 2012 | Not a DT |
| 29 | Contrast between magnetocardiography and electrocardiography for the early diagnosis of coronary artery disease in patients with acute chest pain | Lin LJ, Tang FK, Hua N, and Lu H | 2011 | Different disease |
| 30 | Testing of low-risk patients presenting to the emergency department with chest pain: a scientific statement from the American Heart Association | Amsterdam EA, Kirk JD, Bluemke DA, et al. | 2010 | Not a DT |
| 31 | Non-invasive magnetocardiography for the early diagnosis of coronary artery disease in patients presenting with acute chest pain | Kwon H, Kim K, Lee YH, et al. | 2010 | Different disease |
| 32 | Magnetocardiography based spatiotemporal correlation analysis is superior to conventional ECG analysis for identifying myocardial injury | Goernig M, Liehr M, Tute C, et al. | 2009 | Different disease |
| 33 | Usefulness of magnetocardiogram to detect unstable angina pectoris and non-ST elevation myocardial infarction | Lim HK, Kwon H, Chung N, et al. | 2009 | Different disease |
| 34 | Strain-Encoded Cardiac MR During High-Dose Dobutamine Stress Testing: Comparison to Cine Imaging and to Myocardial Tagging | Korosoglou G, Futterer S, Humpert PM, Riedle N, Lossnitzer D, Hoerig B, et al. | 2009 | Lack of MCG |
| 35 | 冠心病患者的心磁图应用（Clinical application value of magnetocardiography to the patients with coronary heart disease) | Li Y, Jin H, Quan W, Che Z, Yuan R, Shen Y, et al. | 2008 | Not a DT |
| 36 | Reproducibility of quantitative estimate of magnetocardiographic ventricular depolarization and repolarization parameters in healthy subjects and patients with coronary artery disease | Lim HK, Chung N, Kim K, et al. | 2007 | Not a DT |
| 37 | 老年冠心病患者介入治疗前后的心磁图变化（Magnetocardiogram changes after PTCA in old patients with coronary artery diseases) | Du X, Li F, Li Y, Cao J, Li Y, Quan W, et al. | 2007 | Not a DT |
| 38 | Sensitivity and specificity of magnetocardiography, using computerized classification of current density vectors maps, in ischemic patients with normal ECG | Fainzilberg L, Chaikovsky I, Auth-Eisernitz S, Awolin B, | 2007 | Unsatisfactory subjects |

| | | | | |
|---|---|---|---|---|
| | and echocardiogram | Ivaschenko D, and Hailer, B | | |
| 39 | Predictive value of rest magnetocardiography in patients with stable angina | Fenici R and Brisinda D | 2007 | Unsatisfactory reference standard |
| 40 | Integral value of JT interval in magnetocardiography is sensitive to coronary stenosis and improves soon after coronary revascularization | On Kei, Watanabe Shigeyuki, Yamada Satsuki, Takeyasu Norlyuki, Nakagawa Yoshitsugu, Nishina Hidetaka, et al. | 2007 | Not a DT |
| 41 | Application of the TIMI risk score for unstable angina and non-ST elevation acute coronary syndrome to an unselected emergency department chest pain population | Pollack CV Jr, Sites FD, Shofer FS, Sease KL, and Hollander JE | 2006 | Not MCG |
| 42 | 冠心病辅助检查的临床应用进展 (The advancement of clinical applications for auxiliary examination of coronary heart disease) | Niu J | 2006 | Not a DT |
| 43 | 心磁图:临床优势明显,技术仍需完善 (Magnetocardiography: clear clinical advantages, technology still needs improvement) | / | 2006 | Not a DT |
| 44 | 心磁图检查原理及其临床应用 (Principle and Clinical Application of Magnetocardiography) | Wang C, Li W, Zhang J. and Fang P | 2006 | Not a DT |
| 45 | Identification of patients with coronary artery disease using magnetocardiographic signal analysis | Van Leeuwen P, Hailer B, Lange S, and Grönemeyer DH | 2006 | Not a DT of CHD |
| 46 | ACCF/ASNC appropriateness criteria for single-photon emission computed tomography myocardial perfusion imaging (SPECT MPI): a report of the American College of Cardiology Foundation Quality Strategic Directions Committee Appropriateness Criteria Working Group and the American Society of Nuclear Cardiology endorsed by the American Heart Association | Brindis RG, Douglas PS, Hendel RC, et al. | 2005 | Not MCG |
| 47 | 心磁图测量心脏复极时间对冠心病患者心功能预后的评估 (Evaluative role of magenetocardiography on prognosis of cardiac function in patients with coronary heart disease by measuring the time of heart repolarization) | Chen L, Wang Y, Van Leeuwen P, Lange S, and Klein A. | 2005 | Not a DT |
| 48 | The value of magnetocardiography in patients with and without relevant stenoses of the coronary arteries using an unshielded system | Hailer B, Chaikovsky I, Auth-Eisernitz S, Schäfer H, and Van Leeuwen P | 2005 | Unsatisfactory subjects |
| 49 | Myocardial viability evaluation using magnetocardiography in patients with coronary artery disease | Morguet AJ, Behrens S, Kosch O, et al. | 2004 | Not a DT |
| 50 | Qualitative and quantitative description of myocardial ischemia by means of magnetocardiography | Park JW, and Jung F | 2004 | Unsatisfactory reference standard |
| 51 | 冠心病患者的心磁图分析及应用 (Clinical application of magnetocardiography in patients with coronary heart disease) | Li Y, Lu G, Quan W, Shen Y, Yuan R, Du X, et al. | 2004 | Lack of data of diagnostic performance |
| 52 | 心磁图对冠心病患者的诊断价值 (The diagnostic value of magnetocardiography to the patients with coronary heart disease) | Li Y, Lu G, Quan W, Du X, Li F, Yuan R, et al. | 2004 | Unsatisfactory subjects |
| 53 | Detection of coronary artery disease with MCG | Hailer B and Van Leeuwen P | 2004 | Not a DT |
| 54 | Evaluation of magnetocardiography indices in patients with cardiac diseases | Budnyk MM, Kozlovsky VI, Stadnyuk LA, Zahrabova OM, Ryzhenko TM, and Getman TV | 2004 | Not a DT |
| 55 | Age and sex dependent variations in the normal magnetocardiogram compared with changes associated with ischemia | Chen J, Thomson PD, Nolan V, and Clarke J | 2004 | Not a DT |
| 56 | Effects of filtering on computer-aided analysis for detection of chronic ischemic heart disease with unshielded rest magnetocardiography mapping | Fenici R, Brisinda D, and Meloni AM | 2004 | Not a DT |
| 57 | Understanding multimodal fusion imaging | Hoppenrath M | 2004 | Review |

| | | | | |
|---|---|---|---|---|
| 58 | Myocardial viability evaluation using magnetocardiography in patients with coronary artery disease | Morguet AJ, Behrens S, Kosch O, Lange C, Zabel M, Selbig D, et al. | 2004 | Not a DT |
| 59 | 冠心病无创性检查——心磁图 (Magnetocardiography: non-invasive examination for coronary heart disease) | Li W | 2003 | Not a DT |
| 60 | 心磁图在缺血性心脏病的诊断价值探讨 (Preliminary evaluation of the effect of magnetocardiography on diagnosis of ischemic heart disease) | Wang C | 2003 | Unsatisfactory subjects |
| 61 | 心磁图对静态心电图正常的冠心病患者的诊断价值 (Diagnostic value of magnetocardiography in coronary heart disease patients with normal resting electrocardiogram) | Li Y, Lu G, Quan W, Shen Y, and Yuan R | 2003 | Not a DT |
| 62 | First 36-channel magnetocardiographic study of CAD patients in an unshielded laboratory for interventional and intensive cardiac care | Brisinda D, Meloni AM, and Fenici R | 2003 | Unsatisfactory subjects |
| 63 | Coronary artery imaging with real-time navigator three-dimensional turbo-field-echo MR coronary angiography: Initial experience | Bogaert J, Kuzo R, Dymarkowski S, Beckers R, Piessens J, and Rademakers FE | 2003 | Lack of MCG |
| 64 | Comparison of magnetocardiograms acquired in unshielded clinical environment at rest, during and after exercise and in conjunction with myocardial perfusion imaging | Brazdeikis A, Taylor AA, Mahmarian JJ, Xue Y, and Chu CW. | 2002 | Lack of fulltext; not a DT |
| 65 | Computerized classification of patients with coronary artery disease but normal or unspecifically changed ECG and healthy volunteers | Chaikovsky I, Primin M, Nedayvoda I, Vassylyev V, Sosnitsky V, and Steinberg F | 2002 | Lack of fulltext |
| 66 | Evaluation of myocardial ischemia in Kawasaki disease using an isointegral map on magnetocardiogram | Shiono J, Horigome H, Matsui A, Terada Y, Watanabe S, Miyashita T, et al. | 2002 | Different disease |
| 67 | Features of ST segment and T-wave in exercise-induced myocardial ischemia evaluated with multichannel magnetocardiography | Hänninen H, Takala P, Korhonen P, Oikarinen L, Mäkijärvi M, Nenonen J, et al. | 2002 | Not a DT |
| 68 | The continuing search to identify the very-low-risk chest pain patient | Hollander JE | 1999 | Not a DT |
| 69 | Normal and abnormal components in magnetocardiographic maps of a subject with myocardial infarction | Troink G, MacAuley C, Montague TJ, and Horacek BM | 1985 | Not a DT, different disease |
| 70 | Analysis of current source of the heart using isomagnetic and vector arrow maps | Nakaya Y, Sumi M, Saito K, Fujino K, Murakami M, and Mori H | 1984 | Not a DT; different disease |
| 71 | Magnetocardiograms taken inside a shielded room with a superconducting point-contact magnetometer | Cohen D, Edelsack EA, and Zimmerman JE | 1970 | Not a DT |

DT: diagnostic tests; CHD: coronary artery disease; MCG: magnetocardiography

**Table S11 Eliminated studies during the full-text reading**

| No. | Research | Author | Year | Reasons for elimination |
|---|---|---|---|---|
| 1 | 心磁图参数在心电图无特异性改变冠心病患者中应用 (The application of related parameters of MCG in patients of coronary artery disease with normal and unspecialized changed electrocardiogram) | Xu W | 2015 | Overlapping samples with included studies |
| 2 | 高温超导 SQUID 技术在不稳定型心绞痛诊断中的初步研究 (The preliminary investigation of the effectives and possibilities of high temperature superconducting quantum interference device in the diagnosis of unstable angina pectoris) | Wang B | 2014 | Overlapping samples with included studies |

| | | | | |
|---|---|---|---|---|
| 3 | 高温超导量子干涉器在不稳定型心绞痛诊断中的初步研究 (The preliminary investigation of the effectives and possibilities of high temperature superconducting quantum interference device in the diagnosis of unstable angina pectoris) | Wang B, Tang F, Hua N, Di C, Lin L, and Tao Y | 2014 | Overlapping samples with included studies |
| 4 | 四通道高温超导心磁图诊断冠心病的临床初步研究 (A preliminary clinical study on the diagnosis of coronary heart disease with four channel high temperature superconductor MCG) | Yang J | 2013 | Overlapping samples with included studies |
| 5 | Preliminary approach of value of the high-temperature superconducting magnetocardiography in diagnosis of coronary heart disease | Yang J, Tang F, Di C, Zhang C, and Hua N | 2013 | Overlapping samples with included studies |
| 6 | 心磁图对冠心病心肌缺血与高血压心室肥厚的诊断价值 (Effect of magnetocardiography on diagnosis of ischemia in patient with coronary heart disease and left ventricular hypertrophy) | Chen Y | 2010 | Lack of data of diagnostic performance |
| 7 | 心磁图对不稳定型心绞痛的诊断价值 (Diagnostic value of MCG to unstable angina) | Bu L, Tang F, Hua N, Qi Z, Zhang C, and Tang X | 2009 | Unsatisfactory reference standard |
| 8 | 心磁图对冠心病诊断和冠状动脉支架内再狭窄的预测价值 (The predictive value of magnetocardiography for diagnosing coronary heart disease and predicting in-stent restenosis) | Quan W | 2006 | Overlapping samples with included studies |
| 9 | 静息心电图正常的冠心病患者的心磁图分析 | Quan W, Lu G, Li Y, Shen Y, Yuan R, and Qi W | 2006 | Overlapping samples with included studies |
| 10 | Non-invasive resting magnetocardiographic imaging for the rapid detection of ischemia in subjects presenting with chest pain | Tolstrup K, Madsen BE, Ruiz JA, et al. | 2006 | Unsatisfactory reference standard |
| 11 | 冠心病患者 QTd 变异的心磁图研究 (The study of QT dispersion variability in patients with coronary artery disease using magnetocardiography) | Chen L, Van leeuwen P, Lange S, and Klein A | 2004 | Unsatisfactory subjects |
| 12 | 心磁图对常规心电图正常冠心病的诊断价值的初步探讨 (Preliminary approach of diagnostic value of magnetocardiography for coronary artery disease in patients with normal electrocardiogram at rest) | Wang C, Gao R, Hu F, Wei B, Yang Y, You S, et al. | 2004 | Duplicated studies |

### Section 5 Data extraction

| Table S12 Diagnostic 2×2 table | | |
|---|---|---|
| | CAG positive | CAG negative |
| MCG positive | True positives a | False positives b |
| MCG negative | False negatives c | True negatives d |

The main indices include sensitivity $=\frac{a}{a+c}\times 100\%$ and specificity $=\frac{d}{b+d}\times 100\%$. The Youden index, calculated as $\frac{a}{a+c}+\frac{d}{b+d}-1$, evaluates the overall accuracy of the diagnostic test. Other commonly used indices include the false positive rate $=\frac{b}{b+d}$ and false negative rate $=\frac{a}{a+c}$.

**Table S13 Extracted data used for MCG analysis in our study**

| Name | Author | Year | TP | FP | FN | TN | Rest_ecg | Stress_test | Stand_ref |
|---|---|---|---|---|---|---|---|---|---|
| Huang 2019 | Huang | 2019 | 129 | 43 | 3 | 38 | 0 | 0 | 0 |
| Shin 2017 | Shin | 2017 | 39 | 6 | 7 | 44 | 0 | 1 | 1 |
| Wu 2013 | Wu | 2013 | 44 | 7 | 7 | 17 | 0 | 0 | 1 |
| Zhao 2010 | Zhao | 2010 | 216 | 10 | 167 | 100 | 0 | 0 | 0 |
| Zhang 2009 | Zhang | 2009 | 61 | 4 | 8 | 37 | 1 | 0 | 0 |
| Du 2006 | Du | 2006 | 39 | 17 | 7 | 27 | 0 | 0 | 1 |
| Park 2005 | Park | 2005 | 136 | 3 | 7 | 39 | 0 | 0 | 0 |
| Shin 2019 | Shin | 2019 | 54 | 6 | 25 | 117 | 0 | 1 | 1 |
| Kanzaki 2003 | Kanzaki | 2003 | 14 | 2 | 3 | 11 | 1 | 1 | 1 |
| Lin 2015 | Lin | 2015 | 169 | 26 | 21 | 71 | 0 | 0 | 1 |
| Steinberg 2005 | Steinberg | 2005 | 16 | 6 | 3 | 4 | 0 | 0 | 0 |
| Wu 2014 | Wu | 2014 | 31 | 6 | 5 | 13 | 0 | 0 | 1 |
| Chaikovsky 2014 | Chaikovsky | 2014 | 50 | 4 | 4 | 21 | 1 | 0 | 1 |
| Chaikovsky 2017 | Chaikovsky | 2017 | 76 | 6 | 6 | 48 | 0 | 0 | 0 |
| Park 2008 | Park | 2008 | 41 | 19 | 1 | 39 | 0 | 1 | 1 |
| Quan 2006 | Quan | 2006 | 91 | 26 | 49 | 56 | 1 | 0 | 1 |
| Coriasso 2021 | Coriasso | 2021 | 15 | 3 | 12 | 35 | 0 | 0 | |
| Zhao 2019 | Zhao | 2019 | 26 | 29 | 6 | 43 | 0 | 0 | 0 |

Rest_ecg: The patient's resting electrocardiogram (ECG) status, where '1' signifies a normal resting ECG and '0' signifies no specific demand. Stress_test: It indicates whether the patient's MCG was measured under stress, where '1' denotes stress MCG and '0' signifies resting MCG. Stand_ref: It represents the threshold value of the standard reference, where '1' indicating a threshold value of 70% and '0' indicating a threshold value of 50%.

**Section 6 Meta-analysis**

Code for STATA

```
ssc install midas

ssc install mylabels

midas tp fp fn tn, res(sum)

midas tp fp fn tn, id(author year) ms(0.75) ford fors bfor(dss)

midas tp fp fn tn, plot sroc(both)

midas tp fp fn tn, reg(rest_ecg stress_test stand_ref)

midas tp fp fn tn, pubbias
```

**Section 7 Sensitivity analysis**

Funnel Plot

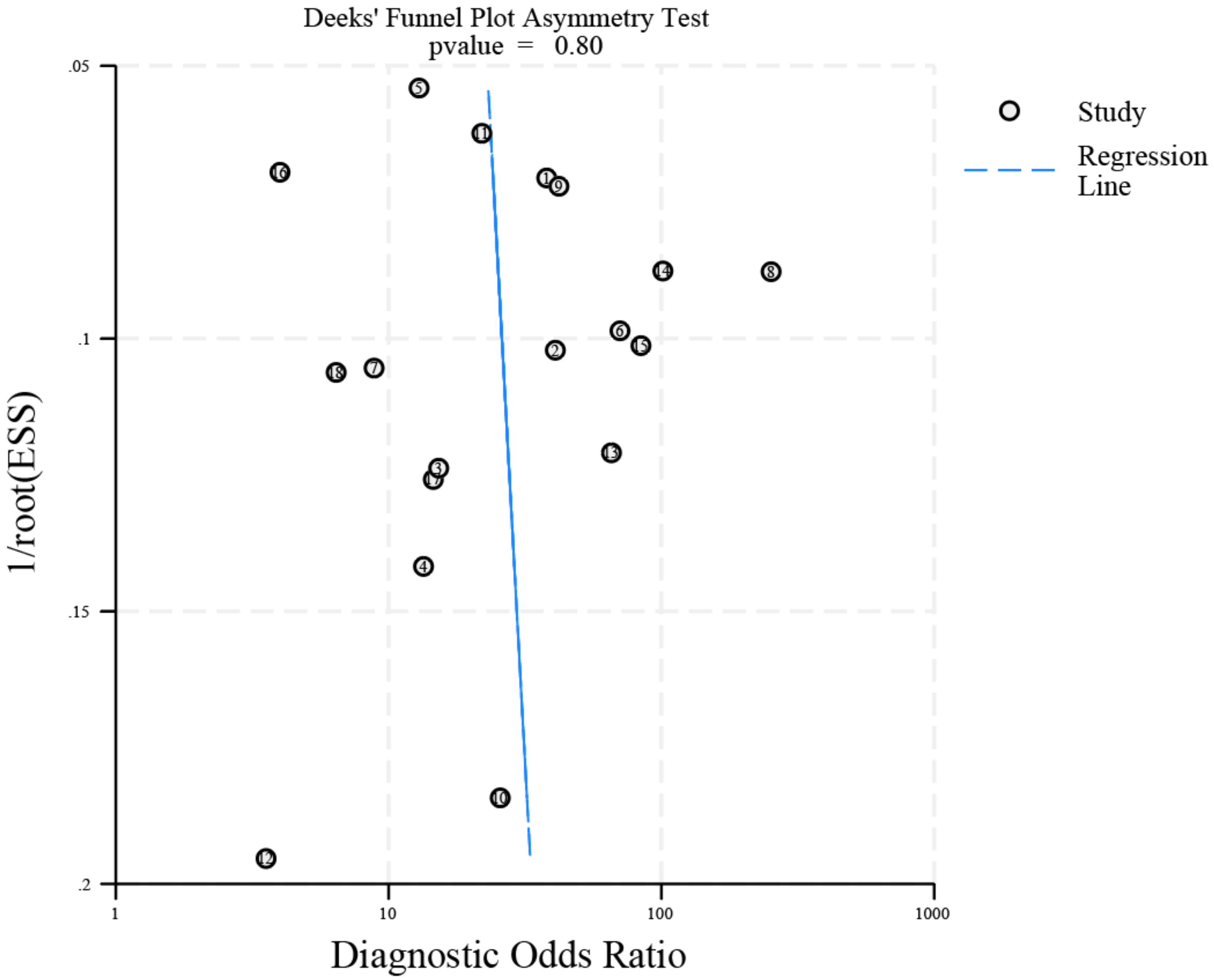

Figure S1 Funnel plot for meta-analysis of MCG. It indicates the absence of significant publication bias

**Section 8 Quality evaluation**

**DOMAIN 1: PATIENT SELECTION**

**Risk of bias: Could the selection of patients have introduced bias?**

Question 1: Was a consecutive or random sample of patients enrolled?

*YES: ‘all patients’, or ‘consecutive patients’.*

Question 2: Was a case-control design avoided?

Question 3: Did the study avoid inappropriate exclusions?

*Exclusion considered inappropriate: patients who are difficult to diagnose based on their clinical presentation.*

*Exclusion considered appropriate: patients who refused entry into the registry, with poor data, with other serious disease, or other unsuitable situations.*

*Risk of bias LOW: If enrolled consecutive or random sample of patients, avoided case-control design, and avoided inappropriate exclusions. Risk of bias Unclear: uncleared exclusions or uncleared recruitment procedures. Risk of bias HIGH: If existed case-control design, discontinuous subjects, or existed inappropriate exclusion.*

**Applicability Concerns: Are there concerns that the included patients and setting do not match the review question?**

*Match the review question: Suspected patients with CHD, using magnetocardiography. High concern: restrictive clinical criteria used at inclusion, leading to a population at higher risk of CHD, or easier to diagnose.*

**DOMAIN 2: INDEX TEST**

**Risk of Bias: Could the conduct or interpretation of the index test have introduced bias?**

Question 1: Were the index test results interpreted without knowledge of the results of the reference standard?

Question 2: If a threshold was used, was it pre-specified?

*Risk of bias LOW: If pre-specified threshold, blinded to the results of coronary angiography. Risk of bias UNCLEAR: If unmentioned blinded design, or unmentioned specified threshold. Risk of bias HIGH: If post-specified threshold or without blinded design.*

**Applicability: Are there concerns that the index test, its conduct, or interpretation differ from the review question?**

*Low concern: same technology, execution, and interpretation. High concern: Existed variation in test technology, execution, or interpretation.*

**DOMAIN 3: REFERENCE STANDARD**

**Risk of Bias: Could the reference standard, its conduct, or its interpretation have introduced bias?**

Question 1: Is the reference standard likely to correctly classify the target condition?

Question 2: Were the reference standard results interpreted without knowledge of the results of the index test?

*Risk of bias LOW: Blinded, with more than 2 independent readers, or at least same experienced reader, with pre-specified criteria. UNCLEAR: unmentioned procedure including unclear readers and unclear blinded design. HIGH: unblinded, without standardized criteria.*

**Applicability: Are there concerns that the target condition as defined by the reference standard does not match the question?**

*Low concern: stenosis more than 50% or 70%.*

**DOMAIN 4: FLOW AND TIMING**

**Risk of Bias: Could the patient flow have introduced bias?**

Question 1: Was there an appropriate interval between index test and reference standard?

*Appropriate interval: Within 48h or other time stated appropriately.*

Question 2: Did all patients receive the same reference standard?

Question 3: Were all patients included in the analysis?

*Risk of Bias LOW: If appropriate interval, same reference standard, and all patients included. Risk of Bias UNCLEAR: Unmentioned interval between index test or unmentioned design of reference standard. HIGH: existed exclusion of patients, or inappropriate interval between index test and reference standard.*

**QUADAS-2 Risk of bias and Applicability concerns Study Reference: Kanzaki 2003**

**DOMAIN 1: PATIENT SELECTION**

**A. Risk of Bias**

*Describe methods of patient selection*: Patients with chest symptoms, who were scheduled for CAG, and healthy volunteers. Recruitment procedure unclear.

- Was a consecutive or random sample of patients enrolled? Unclear
- Was a case-control design avoided? No
- Did the study avoid inappropriate exclusions? Unclear

**Could the selection of patients have introduced bias? Risk: High**

**B. Applicability concerns**

*Describe included patients (prior testing, presentation, intended use of index test and setting):* Patients with chest symptoms and healthy volunteers. All subjects had normal ECG at rest, normal echocardiograms, and no medical devices made of metallic or magnetic material. Aged 38 ± 19 and 61 ± 7 years for control group and CHD patients, respectively. The proportion of male are 85% and 94% for control group and CHD patients, respectively.

**Are there concerns that the included patients and setting do not match the review question? CONCERN: Low**

**DOMAIN 2: INDEX TESTS**

**A. Risk of Bias**

*Describe the index test and how it was conducted and interpreted:* performed with a 64-channel SQUID MCG system installed in a magnetically shielded room. The detectors were positioned just above the anterior chest wall of the subject, who was kept in the supine position. If the maximal QRS integral change during 40 ms ≥ 0.44, the subject was defined as CHD patient.

- Were the index test results interpreted without knowledge of the results of the reference standard? Unclear
- If a threshold was used, was it pre-specified? No

**Could the conduct or interpretation of the index test have introduced bias? Risk: High**

**B. Applicability concerns**

**Are there concerns that the conduce or interpretation of the Index test do not match the review question? CONCERN: Low**

**DOMAIN 3: REFERENCE STANDARD**

**A. Risk of Bias**

*Describe the reference standard and how it was conducted and interpreted*: CAG
*Readers:* Unclear
*Positivity criteria:* > 70% diameter narrowing of one or more major coronary arteries.

- Is the reference standard likely to correctly classify the target condition? Yes
- Were the reference standard results interpreted without knowledge of the results of the index test? Unclear

**Could the conduct or the interpretation of the reference standard have introduced bias?** **Risk: Unclear**

**B. Applicability concerns**

**Are there concerns that the conduct or the interpretation of the reference standard do not match the review question?** **CONCERN: Low**

---

**DOMAIN 4: FLOW AND TIMING**

**A. Risk of Bias**

*Describe any patients who did not receive the index test and/or reference standard or who were excluded from the 2×2 table (refer to flow diagram)*: None.

*Describe the time interval and any interventions between index test and reference standard:* Unclear.

- ➢ Was there an appropriate interval between index test and reference standard? Unclear
- ➢ Did all patients receive the same reference standard? Yes
- ➢ Were all patients included in the analysis? Yes

**Could the flow and timing have introduced bias?** **Risk: Unclear**

**QUADAS-2 Risk of bias and Applicability concerns    Study Reference: Park 2005**

**DOMAIN 1: PATIENT SELECTION**

**A. Risk of Bias**

*Describe methods of patient selection*: Consecutive patients with pains in the retrosternal area and ECG changes indicative of acute ischemic heart disease, without persistent ST-segment elevation, but rather with temporary ST-segment elevation or flat or inverted T waves or even with nonspecific changes at all. Exclusion criteria: Patients with acute myocardial infarction, who were unambiguously diagnosed with a 12-lead surface ECG, hemodynamically unstable patients, and patients who refused entry into the registry.

- Was a consecutive or random sample of patients enrolled? Yes
- Was a case-control design avoided? Yes
- Did the study avoid inappropriate exclusions? Unclear

**Could the selection of patients have introduced bias? Risk: Unclear**

**B. Applicability concerns**

*Describe included patients (prior testing, presentation, intended use of index test and setting):* Aged 67.0 ± 10.7 years. 72 females and 113 males in total.

**Are there concerns that the included patients and setting do not match the review question? CONCERN: Low**

**DOMAIN 2: INDEX TESTS**

**A. Risk of Bias**

*Describe the index test and how it was conducted and interpreted:* 2 independent readers evaluated visually the MCG without prior knowledge of the results of the laboratory results, the 12-lead ECG, the echocardiographic, or the CAG examinations. The detailed procedure and positive criteria not reported.

- Were the index test results interpreted without knowledge of the results of the reference standard? Yes
- If a threshold was used, was it pre-specified? Unclear

**Could the conduct or interpretation of the index test have introduced bias? Risk: Unclear**

**B. Applicability concerns**

**Are there concerns that the conduce or interpretation of the Index test do not match the review question? CONCERN: Low**

**DOMAIN 3: REFERENCE STANDARD**

**A. Risk of Bias**

*Describe the reference standard and how it was conducted and interpreted*: CAG

*Readers:* Unclear

*Criteria for interpretation:* At least one coronary artery branch of first or secondary order revealed a 50% or greater degree of stenosis.

- Is the reference standard likely to correctly classify the target condition? Yes
- Were the reference standard results interpreted without knowledge of the results of the index test? Yes

**Could the conduct or the interpretation of the reference standard have introduced bias? Risk: Low**

**B. Applicability concerns**

**Are there concerns that the conduct or the interpretation of the reference standard do not match the review question? CONCERN: Low**

---

**DOMAIN 4: FLOW AND TIMING**

**A. Risk of Bias**

*Describe any patients who did not receive the index test and/or reference standard or who were excluded from the 2×2 table (refer to flow diagram)*: 264 patients were consecutively recruited and only 185 subjects were evaluated. 8 patients were excluded due to intrinsic signal interference (1 with implanted defibrillator, 2 with metal clips after two bypass operations, 3 with a pacemaker, and 2 had excessive metal after permanent teeth replacement). Another 8 patients were excluded because they became symptom-free after on and no longer needed CAG examination. 63 patients were excluded due to the poor signal to noise ratio of the raw data.

*Describe the time interval and any interventions between index test and reference standard:* Within 36 hours.

- Was there an appropriate interval between index test and reference standard? Yes
- Did all patients receive the same reference standard? Yes
- Were all patients included in the analysis? No

**Could the flow and timing have introduced bias? Risk: High**

| QUADAS-2 Risk of bias and Applicability concerns | Study Reference: Steinberg 2005 |
| --- | --- |

**DOMAIN 1: PATIENT SELECTION**

**A. Risk of Bias**

*Describe methods of patient selection*: Consecutive patients with suspected CHD, who are scheduled for diagnostic coronary artery catheterization. Exclusion criteria: implantable devices, sternal wires, dental artifacts, or otherwise poor data.

- Was a consecutive or random sample of patients enrolled? Unclear
- Was a case-control design avoided? Yes
- Did the study avoid inappropriate exclusions? Unclear

**Could the selection of patients have introduced bias? Risk: Unclear**

**B. Applicability concerns**

*Describe included patients (prior testing, presentation, intended use of index test and setting):* 21 males and 8 females in total, aged from 45 to 83 years.

**Are there concerns that the included patients and setting do not match the review question? CONCERN: Low**

**DOMAIN 2: INDEX TESTS**

**A. Risk of Bias**

*Describe the index test and how it was conducted and interpreted:* Performed using a 9-channel MCG system positioned 3–5 cm above the chest. The subject lies on the examination table. The table can be moved to four different positions, and the MCG is recorded in each of these positions. In each location, the recording is performed for 90 seconds for improvement of the signal/noise ratio. "Common" ECG cycle is used to average each of the 36 MCG tracings over time, then the data are interpolated using first a bivariate surface fitting algorithm to obtain a 2-D color map. Two observers who were blinded to the results of the other tests read the results of MCG. Ischemia on MCG was defined by an algorithm based on three characteristics within the analysis window: the number of poles, the positions and movements of the poles, and the overall stability of the map. The algorithm was preprogrammed into the proprietary analysis software, provided by the manufacturer, and has been submitted for patent. The score of this algorithm ranged from 0 to 100; and greater than 49 was deemed abnormal.

- Were the index test results interpreted without knowledge of the results of the reference standard? Yes
- If a threshold was used, was it pre-specified? Yes

**Could the conduct or interpretation of the index test have introduced bias? Risk: Yes**

**B. Applicability concerns**

**Are there concerns that the conduce or interpretation of the Index test do not match the review question? CONCERN: Low**

**DOMAIN 3: REFERENCE STANDARD**

**A. Risk of Bias**

*Describe the reference standard and how it was conducted and interpreted*: CAG
*Readers:* 2
*Criteria for interpretation:* At least one coronary artery stenosis of 50% was present.

- Is the reference standard likely to correctly classify the target condition? Yes
- Were the reference standard results interpreted without knowledge of the results of the index test? Unclear

**Could the conduct or the interpretation of the reference standard have introduced bias? Risk: Unclear**

**B. Applicability concerns**

**Are there concerns that the conduct or the interpretation of the reference standard do not match the review question? CONCERN: Low**

---

**DOMAIN 4: FLOW AND TIMING**

**A. Risk of Bias**

*Describe any patients who did not receive the index test and/or reference standard or who were excluded from the 2 ×2 table (refer to flow diagram)*: None.
*Describe the time interval and any interventions between index test and reference standard:* Within 24 hours.

- Was there an appropriate interval between index test and reference standard? Yes
- Did all patients receive the same reference standard? Yes
- Were all patients included in the analysis? Yes

**Could the flow and timing have introduced bias? Risk: Low**

**QUADAS-2 Risk of bias and Applicability concerns Study Reference: Du 2006**

**DOMAIN 1: PATIENT SELECTION**

**A. Risk of Bias**

*Describe methods of patient selection*: Consecutive patients with chest pain. Exclusion criteria: patients with heart failure, electrolyte imbalance, atrial fibrillation, bundle branch block, and left ventricular hypertrophy confirmed by echocardiography, age < 26 years old.

- Was a consecutive or random sample of patients enrolled? Yes
- Was a case-control design avoided? Yes
- Did the study avoid inappropriate exclusions? Unclear

**Could the selection of patients have introduced bias? Risk: Unclear**

**B. Applicability concerns**

*Describe included patients (prior testing, presentation, intended use of index test and setting):* 90 subjects in total, with 61 males and 29 females.

**Are there concerns that the included patients and setting do not match the review question? CONCERN: Low**

**DOMAIN 2: INDEX TESTS**

**A. Risk of Bias**

*Describe the index test and how it was conducted and interpreted:* Sensors were placed 1-2 cm above the chest and its recording cover an area of 20 × 20 cm. The subject lied on the examination bed with all magnetic, electronic and metallic objects removed. The patients are diagnosed according to the ratio of abnormal maps RAM in the S-T interval. The cut-off value decided by ROC curve not reported.

- Were the index test results interpreted without knowledge of the results of the reference standard? Unclear
- If a threshold was used, was it pre-specified? Unclear

**Could the conduct or interpretation of the index test have introduced bias? Risk: Unclear**

**B. Applicability concerns**

**Are there concerns that the conduce or interpretation of the Index test do not match the review question? CONCERN: Low**

**DOMAIN 3: REFERENCE STANDARD**

**A. Risk of Bias**

*Describe the reference standard and how it was conducted and interpreted*: CAG

*Readers:* Unclear

*Criteria for interpretation:* At least one coronary artery stenosis of 70% was present.

- Is the reference standard likely to correctly classify the target condition? Yes
- Were the reference standard results interpreted without knowledge of the results of the index test? Unclear

**Could the conduct or the interpretation of the reference standard have introduced bias? Risk: Unclear**

**B. Applicability concerns**

**Are there concerns that the conduct or the interpretation of the reference standard do not match the review question? CONCERN: Low**

---

**DOMAIN 4: FLOW AND TIMING**

**A. Risk of Bias**

*Describe any patients who did not receive the index test and/or reference standard or who were excluded from the 2 ×2 table (refer to flow diagram)*: None.

*Describe the time interval and any interventions between index test and reference standard:* Unclear

- Was there an appropriate interval between index test and reference standard? Unclear
- Did all patients receive the same reference standard? Yes
- Were all patients included in the analysis? Yes

**Could the flow and timing have introduced bias? Risk: Unclear**

**QUADAS-2 Risk of bias and Applicability concerns Study Reference: Quan 2006**

**DOMAIN 1: PATIENT SELECTION**

**A. Risk of Bias**

*Describe methods of patient selection*: Consecutive patients with chest pain who exhibit normal findings in echocardiography, chest X-rays, and resting electrocardiogram (ECG) recordings. Exclusion criteria: Patients with heart failure, bundle branch block, atrial fibrillation. Patients with cardiomyopathy, valvular heart disease, ventricular hypertrophy, and left ventricular dysfunction confirmed by Doppler ultrasound.

- Was a consecutive or random sample of patients enrolled? Yes
- Was a case-control design avoided? Yes
- Did the study avoid inappropriate exclusions? Unclear

**Could the selection of patients have introduced bias? Risk: Unclear**

**B. Applicability concerns**

*Describe included patients (prior testing, presentation, intended use of index test and setting):* Inclusion and exclusion criteria see above. Aged from 37 to 85 years and from 43 to 78 years for CHD group and non-CHD group, respectively.

**Are there concerns that the included patients and setting do not match the review question? CONCERN: Low**

**DOMAIN 2: INDEX TESTS**

**A. Risk of Bias**

*Describe the index test and how it was conducted and interpreted:* MCG is recorded in an unshielded room at a 5 × 5 rectangular grid and separated from each other by 4 cm. MCG of all the measuring points during ST segment is utilized to reconstruct a current density figure. The patient with the value of complex ventricular excitation index (CVEI) between -100 to 0 is diagnosed as CHD.

- Were the index test results interpreted without knowledge of the results of the reference standard? Unclear
- If a threshold was used, was it pre-specified? YES

**Could the conduct or interpretation of the index test have introduced bias? Risk: Unclear**

**B. Applicability concerns**

**Are there concerns that the conduce or interpretation of the Index test do not match the review question? CONCERN: Low**

**DOMAIN 3: REFERENCE STANDARD**

**A. Risk of Bias**

*Describe the reference standard and how it was conducted and interpreted*: CAG
*Readers:* Unclear
*Criteria for interpretation:* At least one main coronary artery stenosis of >= 70%.

- Is the reference standard likely to correctly classify the target condition? Yes

- Were the reference standard results interpreted without knowledge of the results of the index test? Unclear

**Could the conduct or the interpretation of the reference standard have introduced bias? Risk: Low**

**B. Applicability concerns**

**Are there concerns that the conduct or the interpretation of the reference standard do not match the review question? CONCERN: Low**

---

**DOMAIN 4: FLOW AND TIMING**

**A. Risk of Bias**

| |
|---|
| *Describe any patients who did not receive the index test and/or reference standard or who were excluded from the 2×2 table (refer to flow diagram)*: None.<br>*Describe the time interval and any interventions between index test and reference standard:* Unclear. |

- Was there an appropriate interval between index test and reference standard? Unclear
- Did all patients receive the same reference standard? Yes
- Were all patients included in the analysis? Yes

**Could the flow and timing have introduced bias? Risk: Unclear**

---

**QUADAS-2 Risk of bias and Applicability concerns** **Study Reference: Park 2008**

**DOMAIN 1: PATIENT SELECTION**

**A. Risk of Bias**

*Describe methods of patient selection*: Consecutive patients with stable angina pectoris or exertional dyspnea and an intermediate pre-test probability (10–90%) for CHD, who were referred to Hoyerswerda Hospital for the first CAG. Exclusion criteria included: 1. Age <18 years 2. Acute coronary syndromes 3. Low (<10%) or high (>90%) pre-test; probability for CHD 4. History of percutaneous coronary intervention (PCI) 5. History of coronary bypass surgery 6. Hypertrophic obstructive cardiomyopathy (HOCM) 7. Hypertrophic nonobstructive cardiomyopathy (HNOCM) 8. Dilated cardiomyopathy (DCM) 9. Aortic valve stenosis >◦2 10. Mitral valve stenosis >◦2 11. Complete bundle branch block Glaucoma 12. Hypovolemia 13. History of contrast agent allergy 14. Creatinine >135 mmol/l 15. Pregnancy

- Was a consecutive or random sample of patients enrolled? Unclear
- Was a case-control design avoided? Yes
- Did the study avoid inappropriate exclusions? Unclear

**Could the selection of patients have introduced bias? Risk: Unclear**

**B. Applicability concerns**

*Describe included patients (prior testing, presentation, intended use of index test and setting):* Inclusion and exclusion criteria see above. There are 49 males and 51 females, with a mean age of 65.1 ± 10.1 years old. Mean value of height and body weight are 168 ± 9.7 cm and 81.8 ± 15.5 kg, respectively.

**Are there concerns that the included patients and setting do not match the review question? CONCERN: Low**

**DOMAIN 2: INDEX TESTS**

**A. Risk of Bias**

*Describe the index test and how it was conducted and interpreted:* Performed using a 55 channel MCG system (intersensor spacing 3.2 cm covering a circular area of about 23 cm) in a magnetically shielded room. The patients were examined at rest and during the dobutamine–atropine stress until age-adjusted submaximal heart rate was reached. The epicardial current distribution was calculated and reconstructed from the magnetic field data and superposed on a virtual heart model (Software: AtB, Pescara, Italy and Biomagnetik, Cologne, Germany). A head-to-head visual comparison is used for diagnosing CHD. Patients with a significant reduction of epicardial current density and current strength at the timepoint of maximal strength of QRS-complex (QRSmax) during stress is diagnosed as CHD.

- Were the index test results interpreted without knowledge of the results of the reference standard? Yes
- If a threshold was used, was it pre-specified? Unclear

**Could the conduct or interpretation of the index test have introduced bias? Risk: Unclear**

**B. Applicability concerns**

**Are there concerns that the conduce or interpretation of the Index test do not match the review question? CONCERN: Low**

**DOMAIN 3: REFERENCE STANDARD**

**A. Risk of Bias**

*Describe the reference standard and how it was conducted and interpreted*: CAG
*Readers:* 1
*Criteria for interpretation:* Lesions with a diameter reduction of >= 70%.

- Is the reference standard likely to correctly classify the target condition? Yes
- Were the reference standard results interpreted without knowledge of the results of the index test? Unclear

**Could the conduct or the interpretation of the reference standard have introduced bias? Risk: Unclear**

**B. Applicability concerns**

**Are there concerns that the conduct or the interpretation of the reference standard do not match the review question? CONCERN: Low**

---

**DOMAIN 4: FLOW AND TIMING**

**A. Risk of Bias**

*Describe any patients who did not receive the index test and/or reference standard or who were excluded from the 2×2 table (refer to flow diagram)*: None.
*Describe the time interval and any interventions between index test and reference standard:* Within 24 hours.

- Was there an appropriate interval between index test and reference standard? Yes
- Did all patients receive the same reference standard? Yes
- Were all patients included in the analysis? Yes

**Could the flow and timing have introduced bias? Risk: Low**

**QUADAS-2 Risk of bias and Applicability concerns        Study Reference: Zhang 2009**

**DOMAIN 1: PATIENT SELECTION**

**A. Risk of Bias**

*Describe methods of patient selection*: Consecutive patients with chest pain or chest discomfort. Inclusion criteria included: 1. Routine 12-lead resting ECG shows normal findings or exhibits non-specific changes. 2. Creatine kinase-MB isoenzyme (CK-MB) is less than 2 times the normal range. 3. Troponin I ≤0.16 ng/L 4. aged from 50 - 70 years old. Exclusion criteria included: 1. Acute and old myocardial infarction. 2. Primary and secondary hypertension. 3. Clinically significant valve disease. 4. Diabetes mellitus 5. Atrioventricular block, bundle branch block, intraventricular block, and post-pacemaker implantation. 6. LVEF <= 50%. 7. Other situations that the researchers deemed unsuitable for inclusion.

- ➢ Was a consecutive or random sample of patients enrolled? Yes
- ➢ Was a case-control design avoided? Yes
- ➢ Did the study avoid inappropriate exclusions? Unclear

**Could the selection of patients have introduced bias? Risk: Unclear**

**B. Applicability concerns**

*Describe included patients (prior testing, presentation, intended use of index test and setting):* See above. No details in the patients' characteristics reported.

**Are there concerns that the included patients and setting do not match the review question? CONCERN: Low**

**DOMAIN 2: INDEX TESTS**

**A. Risk of Bias**

*Describe the index test and how it was conducted and interpreted:* Performed using a 9-channel MCG system in an unshielded room. All magnetic, electronic and metallic objects were removed. The sensors were placed at 1-2 cm above the chest and recorded 36 points in total. The MCG signals were sent to the computer and then analyzed. Patients were diagnosed as CHD if at least 3 out of the following 7 indicators were found to be abnormal: (1) Magnetic dipole standard integral, (2) Magnetic dipole quantitative integral, (3) Magnetic dipole standard scoring, (4) Magnetic dipole angle range, (5) Magnetic dipole angle change, (6) Magnetic dipole distance change, (7) Positive and negative extreme value change ratio. The threshold of these parameters not reported.

- ➢ Were the index test results interpreted without knowledge of the results of the reference standard? Unclear
- ➢ If a threshold was used, was it pre-specified? Unclear

**Could the conduct or interpretation of the index test have introduced bias? Risk: Unclear**

**B. Applicability concerns**

**Are there concerns that the conduce or interpretation of the Index test do not match the review question? CONCERN: Low**

**DOMAIN 3: REFERENCE STANDARD**

**A. Risk of Bias**

*Describe the reference standard and how it was conducted and interpreted*: CAG

*Readers:* Unclear

*Criteria for interpretation:* At least one major coronary artery or its major branch has ≥50% diameter stenosis.

- Is the reference standard likely to correctly classify the target condition? Yes
- Were the reference standard results interpreted without knowledge of the results of the index test? Unclear

**Could the conduct or the interpretation of the reference standard have introduced bias? Risk: Unclear**

**B. Applicability concerns**

**Are there concerns that the conduct or the interpretation of the reference standard do not match the review question? CONCERN: Low**

**DOMAIN 4: FLOW AND TIMING**

**A. Risk of Bias**

*Describe any patients who did not receive the index test and/or reference standard or who were excluded from the 2×2 table (refer to flow diagram)*: None.

*Describe the time interval and any interventions between index test and reference standard:* Within 24 hours.

- Was there an appropriate interval between index test and reference standard? Yes
- Did all patients receive the same reference standard? Yes
- Were all patients included in the analysis? Yes

**Could the flow and timing have introduced bias? Risk: Low**

**QUADAS-2 Risk of bias and Applicability concerns Study Reference: Zhao 2010**

**DOMAIN 1: PATIENT SELECTION**

**A. Risk of Bias**

*Describe methods of patient selection*: Consecutive patients with suspected CHD exhibiting chest pain. Exclusion criteria included: 1. Severe hypertension ( > 180/110 mmHg) 2. Complex arrhythmias such as frequent atrial premature beats, ventricular premature beats, and complete bundle branch block. 3. patients with severe lung disease, chest deformity, or those who have undergone surgery. 4. Patients with acute myocardial infarction of less than 3 months. 5. Patients with ventricular hypertrophy or dilated cardiomyopathy confirmed by echocardiography.

- Was a consecutive or random sample of patients enrolled? Yes
- Was a case-control design avoided? Yes
- Did the study avoid inappropriate exclusions? Unclear

**Could the selection of patients have introduced bias? Risk: Unclear**

**B. Applicability concerns**

*Describe included patients (prior testing, presentation, intended use of index test and setting):* The CHD group consists of 383 subjects, including 253 males and 130 females. The mean value of the age is 59.56 ± 9.60 years. The non-CHD group consists of 110 subjects, including 58 males and 52 females. The mean value of the age is 51.15 ± 8.60 years.

**Are there concerns that the included patients and setting do not match the review question? CONCERN: Low**

**DOMAIN 2: INDEX TESTS**

**A. Risk of Bias**

*Describe the index test and how it was conducted and interpreted:* Performed using 9-channel MCG system, and all sensors are aligned as a rectangle of 3 × 3, with the distance of 4 cm between neighboring sensors. All magnetic, electronic and metallic objects were removed. The system is placed at 4 positions to record MCG at 36 points in total. In each location, the recording is performed for 90 seconds. Patients were diagnosed as CHD if at least 3 out of the following 7 indicators were found to be abnormal: 1. pre-peak repolarization angle < -120 or > -25. 2. pre-peak repolarization trajectory ≥4.3. 3. pre-peak repolarization angular deviation ≥ 0.5. 4. post-peak repolarization angle < -110 or > -22. 5. post-peak repolarization trajectory ≥ 3.7. 6. post-peak repolarization angular deviation ≥ 0.45. 7. prepost angle change < -35 or > -12.

- Were the index test results interpreted without knowledge of the results of the reference standard? Unclear
- If a threshold was used, was it pre-specified? Unclear

**Could the conduct or interpretation of the index test have introduced bias? Risk: Unclear**

**B. Applicability concerns**

**Are there concerns that the conduce or interpretation of the Index test do not match the review question? CONCERN: Low**

**DOMAIN 3: REFERENCE STANDARD**

**A. Risk of Bias**

*Describe the reference standard and how it was conducted and interpreted*: CAG
*Readers:* Unclear
*Criteria for interpretation:* At least one major branch of coronary artery stenosis ≥50%.

- Is the reference standard likely to correctly classify the target condition? Yes
- Were the reference standard results interpreted without knowledge of the results of the index test? Unclear

**Could the conduct or the interpretation of the reference standard have introduced bias? Risk: Unclear**

**B. Applicability concerns**

**Are there concerns that the conduct or the interpretation of the reference standard do not match the review question? CONCERN: Low**

---

**DOMAIN 4: FLOW AND TIMING**

**A. Risk of Bias**

*Describe any patients who did not receive the index test and/or reference standard or who were excluded from the 2×2 table (refer to flow diagram)*: None.
*Describe the time interval and any interventions between index test and reference standard:* Unclear

- Was there an appropriate interval between index test and reference standard? Unclear
- Did all patients receive the same reference standard? Yes
- Were all patients included in the analysis? Yes

**Could the flow and timing have introduced bias? Risk: Unclear**

**QUADAS-2 Risk of bias and Applicability concerns Study Reference: Wu 2013**

**DOMAIN 1: PATIENT SELECTION**

**A. Risk of Bias**

*Describe methods of patient selection*: Consecutive patients with suspected CHD. Exclusion criteria: significant arrhythmias, known MI history or Q wave on surface 12-lead ECG, unstable angina pectoris, significant valvular heart disease, metallic prosthesis (including pacemaker and implantable cardioverter defibrillator).

- Was a consecutive or random sample of patients enrolled? Yes
- Was a case-control design avoided? Yes
- Did the study avoid inappropriate exclusions? Unclear

**Could the selection of patients have introduced bias? Risk: Unclear**

**B. Applicability concerns**

*Describe included patients (prior testing, presentation, intended use of index test and setting):* The mean age of the patients is 64 ± 10 years old. 67 males and 8 females in total.

**Are there concerns that the included patients and setting do not match the review question? CONCERN: Low**

**DOMAIN 2: INDEX TESTS**

**A. Risk of Bias**

*Describe the index test and how it was conducted and interpreted:* Performed using a 64-channel SQUID system in a magnetically shielded room. The MCG signals were digitally recorded for 100s at a sampling rate of 500Hz, with the patient in the supine position and the SQUID's 2-D arrayed sensors positioned close to, but not in contact with, the left chest wall. After baseline correction, data were averaged using R-peaks to obtain a time-averaged 1-period MCG signal. MCG signals during the QT interval was used for the construction of the QT contour map. QTc dispersion and SI-QTc are derived from the QT contour map to represent the myocardial repolarization heterogeneity. Patients with the QTc dispersion ≥79ms or SI-QTc ≥9ms are diagnosed as CHD.

- Were the index test results interpreted without knowledge of the results of the reference standard? Unclear
- If a threshold was used, was it pre-specified? No

**Could the conduct or interpretation of the index test have introduced bias? Risk: High**

**B. Applicability concerns**

**Are there concerns that the conduce or interpretation of the Index test do not match the review question? CONCERN: Low**

**DOMAIN 3: REFERENCE STANDARD**

**A. Risk of Bias**

*Describe the reference standard and how it was conducted and interpreted*: CAG

*Readers: /*

*Criteria for interpretation:* Angiographic maximum lesions ≥50% luminal stenosis in the left main (LM), or ≥70% in at least one of the primary coronary arteries and their major branches.

- Is the reference standard likely to correctly classify the target condition? Yes
- Were the reference standard results interpreted without knowledge of the results of the index test? Unclear

**Could the conduct or the interpretation of the reference standard have introduced bias? Risk: Unclear**

**B. Applicability concerns**

**Are there concerns that the conduct or the interpretation of the reference standard do not match the review question? CONCERN: Low**

**DOMAIN 4: FLOW AND TIMING**

**A. Risk of Bias**

| |
|---|
| *Describe any patients who did not receive the index test and/or reference standard or who were excluded from the 2×2 table (refer to flow diagram)*: None.<br>*Describe the time interval and any interventions between index test and reference standard:* within 1 month. |

- Was there an appropriate interval between index test and reference standard? Yes
- Did all patients receive the same reference standard? Yes
- Were all patients included in the analysis? Yes

**Could the flow and timing have introduced bias? Risk: Low**

**QUADAS-2 Risk of bias and Applicability concerns** **Study Reference: Chaikovsky 2014**

**DOMAIN 1: PATIENT SELECTION**

**A. Risk of Bias**

*Describe methods of patient selection*: Patients with chest pain. Exclusion. Exclusion criteria: Patients with previous myocardial infarction, atrial fibrillation and flutter, pacemaker, heart failure, hypertensive disease of stage III, renal and hepatic failure, obstructive respiratory diseases, febrile states, or oncological diseases were excluded. Further exclusion criteria were abnormalities in 12-lead ECG at rest (abnormal Q-waves, ST depression/elevation, negative T-waves in more than two leads, complete bundle branch block, signs of the left ventricular hypertrophy) as well as abnormalities in rest echocardiogram (segmental and global disturbances of contractility, enlargement of hear chamber, valvular heart disease). No details on patient recruitment.

- Was a consecutive or random sample of patients enrolled? Unclear
- Was a case-control design avoided? Yes
- Did the study avoid inappropriate exclusions? Unclear

**Could the selection of patients have introduced bias?** **Risk: Unclear**

**B. Applicability concerns**

*Describe included patients (prior testing, presentation, intended use of index test and setting):* The CHD group consists of 54 subjects, including 48 males and 6 females. The mean value of the age is 56 ± 10 years. The non-CHD group consists of 25 subjects, including 20 males and 5 females. The mean value of the age is 54 ± 12 years.

**Are there concerns that the included patients and setting do not match the review question?** **CONCERN: Low**

**DOMAIN 2: INDEX TESTS**

**A. Risk of Bias**

*Describe the index test and how it was conducted and interpreted:* MCG was conducted using a 7-channel stationary MCG system in an unshielded room. Patients were positioned on a movable platform, and sensors were placed at nine locations over the precordial region. The magnetic field registration lasted for 30 seconds to 1 minute at each location, while simultaneously recording a two-lead ECG. Trained nurses performed the MCG registration, and the data were filtered and processed. Averaged MCG curves were obtained and analyzed, with reference points marked based on the QRS complex and ST-T interval. The signal-to-noise ratio was suitable for analysis, and the PQRST reference points were well differentiated. Written informed consent was obtained from all patients. Patient is diagnosed as CHD if the complex index CI >= 10.

- Were the index test results interpreted without knowledge of the results of the reference standard? Unclear
- If a threshold was used, was it pre-specified? No

**Could the conduct or interpretation of the index test have introduced bias?** **Risk: High**

**B. Applicability concerns**

**Are there concerns that the conduce or interpretation of the Index test do not match the review question?** **CONCERN: Low**

**DOMAIN 3: REFERENCE STANDARD**

**A. Risk of Bias**

*Describe the reference standard and how it was conducted and interpreted*: CAG
*Readers:* 2
*Criteria for interpretation:* Detected stenosis of 70% or more in at least one of the main coronary arteries.

- Is the reference standard likely to correctly classify the target condition? Yes
- Were the reference standard results interpreted without knowledge of the results of the index test? Unclear

**Could the conduct or the interpretation of the reference standard have introduced bias? Risk: Unclear**

**B. Applicability concerns**

**Are there concerns that the conduct or the interpretation of the reference standard do not match the review question? CONCERN: Low**

**DOMAIN 4: FLOW AND TIMING**

**A. Risk of Bias**

*Describe any patients who did not receive the index test and/or reference standard or who were excluded from the 2×2 table (refer to flow diagram)*: None.
*Describe the time interval and any interventions between index test and reference standard:* 24 – 48 hours.

- Was there an appropriate interval between index test and reference standard? Unclear
- Did all patients receive the same reference standard? Yes
- Were all patients included in the analysis? Yes

**Could the flow and timing have introduced bias? Risk: Unclear**

**QUADAS-2 Risk of bias and Applicability concerns** **Study Reference: Wu 2014**

**DOMAIN 1: PATIENT SELECTION**

**A. Risk of Bias**

*Describe methods of patient selection*: Consecutive patients with suspected or known stable CAD referred for stress MPI and CAG. Inclusion criteria included: 1. typical/atypical chest pain or ischemic equivalents (eg. dyspnea). 2. an interpretable baseline electrocardiography (ECG) and in sinus rhythm. 3. at least intermediate pretest CHD likelihood. 4. preserved left ventricular (LV) ejection fraction (EF) (>= 50% by 2D echocardiography) and wall motion. Exclusion criteria: significant arrhythmias, recent (< 6 weeks) myocardial infarction, unstable angina pectoris, Q-wave on 12-lead ECG, and metallic prosthesis (including pacemaker and implantable cardioverter-defibrillator).

- Was a consecutive or random sample of patients enrolled? Yes
- Was a case-control design avoided? Yes
- Did the study avoid inappropriate exclusions? Unclear

**Could the selection of patients have introduced bias? Risk: Unclear**

**B. Applicability concerns**

*Describe included patients (prior testing, presentation, intended use of index test and setting):* For CHD group and non-CHD group, the mean values of age are 64 ± 10 and 65 ± 9 years old, respectively. There is no significant difference in age, gender within these 2 groups.

**Are there concerns that the included patients and setting do not match the review question? CONCERN: Low**

**DOMAIN 2: INDEX TESTS**

**A. Risk of Bias**

*Describe the index test and how it was conducted and interpreted:* Performed using a 64-channel SQUID system in a magnetically shielded room. The MCG signals were digitally recorded for 100s at a sampling rate of 500Hz, with the patient in the supine position and the SQUID's 2-D arrayed sensors positioned close to, but not in contact with, the left chest wall. After baseline correction, data were averaged using R-peaks to obtain a time-averaged 1-period MCG signal. MCG signals during the QT interval was used for the construction of the QT contour map. QTc dispersion and SI-QTc are derived from the QT contour map to represent the myocardial repolarization heterogeneity. Patients with the QTc dispersion ≥79ms or SI-QTc ≥9ms are diagnosed as CHD.

- Were the index test results interpreted without knowledge of the results of the reference standard? Unclear
- If a threshold was used, was it pre-specified? No

**Could the conduct or interpretation of the index test have introduced bias? Risk: High**

**B. Applicability concerns**

**Are there concerns that the conduce or interpretation of the Index test do not match the review question? CONCERN: Low**

**DOMAIN 3: REFERENCE STANDARD**

**A. Risk of Bias**

*Describe the reference standard and how it was conducted and interpreted*: CAG

*Readers: /*

*Criteria for interpretation:* Angiographic maximum lesions ≥50% luminal stenosis in the left main (LM), or ≥70% in at least one of the primary coronary arteries and their major branches.

- Is the reference standard likely to correctly classify the target condition? Yes
- Were the reference standard results interpreted without knowledge of the results of the index test? Unclear

**Could the conduct or the interpretation of the reference standard have introduced bias? Risk: Unclear**

**B. Applicability concerns**

**Are there concerns that the conduct or the interpretation of the reference standard do not match the review question? CONCERN: Low**

---

**DOMAIN 4: FLOW AND TIMING**

**A. Risk of Bias**

*Describe any patients who did not receive the index test and/or reference standard or who were excluded from the 2×2 table (refer to flow diagram)*: Patient with suspected CHD

*Describe the time interval and any interventions between index test and reference standard:* Within 3 months.

- Was there an appropriate interval between index test and reference standard? Yes
- Did all patients receive the same reference standard? Yes
- Were all patients included in the analysis? Yes

**Could the flow and timing have introduced bias? Risk: Low**

**QUADAS-2 Risk of bias and Applicability concerns Study Reference: Lin 2015**

**DOMAIN 1: PATIENT SELECTION**

**A. Risk of Bias**

*Describe methods of patient selection*: Consecutive patients with chest pain or chest discomfort. Exclusion criteria: 1. patients with suspected variant angina, persistent ST segment elevation, bundle branch block, arrhythmias. 2. patients with left ventricular hypertrophy confirmed by echocardiography.

- Was a consecutive or random sample of patients enrolled? Yes
- Was a case-control design avoided? Yes
- Did the study avoid inappropriate exclusions? Unclear

**Could the selection of patients have introduced bias? Risk: Unclear**

**B. Applicability concerns**

*Describe included patients (prior testing, presentation, intended use of index test and setting):* See above. No details in the patients' characteristics reported.

**Are there concerns that the included patients and setting do not match the review question? CONCERN: Low**

**DOMAIN 2: INDEX TESTS**

**A. Risk of Bias**

*Describe the index test and how it was conducted and interpreted:* Performed using a 9-channel MCG system in the magnetically shielded room. Then the MCG signals were used to calculate seven parameters. Patients were diagnosed as CHD if at least 3 out of the following 7 indicators were found to be abnormal: 1. pre-peak repolarization angle < -120 or > -25. 2. pre-peak repolarization trajectory ≥4.3. 3. pre-peak repolarization angular deviation ≥ 0.5. 4. post-peak repolarization angle < -110 or > -22. 5. post-peak repolarization trajectory ≥ 3.7. 6. post-peak repolarization angular deviation ≥ 0.45. 7. prepost angle change < -35 or > -12.

- Were the index test results interpreted without knowledge of the results of the reference standard? Unclear
- If a threshold was used, was it pre-specified? Unclear

**Could the conduct or interpretation of the index test have introduced bias? Risk: Unclear**

**B. Applicability concerns**

**Are there concerns that the conduce or interpretation of the Index test do not match the review question? CONCERN: Low**

**DOMAIN 3: REFERENCE STANDARD**

**A. Risk of Bias**

*Describe the reference standard and how it was conducted and interpreted*: CAG

*Readers:* Unclear

*Criteria for interpretation:* At least 1 stenosis >= 70% in the three main coronary arteries and their branches.

- Is the reference standard likely to correctly classify the target condition? Yes
- Were the reference standard results interpreted without knowledge of the results of the index test? Unclear

**Could the conduct or the interpretation of the reference standard have introduced bias? Risk: Unclear**

**B. Applicability concerns**

**Are there concerns that the conduct or the interpretation of the reference standard do not match the review question? CONCERN: Low**

---

**DOMAIN 4: FLOW AND TIMING**

**A. Risk of Bias**

*Describe any patients who did not receive the index test and/or reference standard or who were excluded from the 2×2 table (refer to flow diagram)*: None.

*Describe the time interval and any interventions between index test and reference standard:* Unclear.

- Was there an appropriate interval between index test and reference standard? Unclear
- Did all patients receive the same reference standard? Yes
- Were all patients included in the analysis? Yes

**Could the flow and timing have introduced bias? Risk: Unclear**

**QUADAS-2 Risk of bias and Applicability concerns Study Reference: Chaikovsky 2017**

**DOMAIN 1: PATIENT SELECTION**

**A. Risk of Bias**

*Describe methods of patient selection*: Patients, who exhibiting symptoms of chest pain, without a history of myocardial infarction. Exclusion criteria and recruitment procedure undetailed.

- Was a consecutive or random sample of patients enrolled? Unclear
- Was a case-control design avoided? Yes
- Did the study avoid inappropriate exclusions? Unclear

**Could the selection of patients have introduced bias? Risk: Unclear**

**B. Applicability concerns**

*Describe included patients (prior testing, presentation, intended use of index test and setting):* 82 and 54 subjects for CHD group and non-CHD group, respectively. No details in patients' characteristics reported.

**Are there concerns that the included patients and setting do not match the review question? CONCERN: Low**

**DOMAIN 2: INDEX TESTS**

**A. Risk of Bias**

*Describe the index test and how it was conducted and interpreted:* The magnetocardiography recordings were obtained from 36 positions during rest, and CDV maps were generated during the ST-T interval. Each CDV map element was characterized by brightness, representing the current density, and the angle of the current density vector at that point. This resulted in the calculation of 32 features for each map. A binary k-NN classifier employing different distance metrics (Cityblock, Mahalanobis, Chebyshev, Euclidean) was utilized to classify the patient into the relevant categories.

- Were the index test results interpreted without knowledge of the results of the reference standard? Unclear
- If a threshold was used, was it pre-specified? Unclear

**Could the conduct or interpretation of the index test have introduced bias? Risk: Unclear**

**B. Applicability concerns**

**Are there concerns that the conduce or interpretation of the Index test do not match the review question? CONCERN: Low**

**DOMAIN 3: REFERENCE STANDARD**

**A. Risk of Bias**

*Describe the reference standard and how it was conducted and interpreted*: CAG
*Readers:* Unclear
*Criteria for interpretation:* At least 50% stenosis in at least one of the main coronary arteries.

- Is the reference standard likely to correctly classify the target condition? Yes
- Were the reference standard results interpreted without knowledge of the results of the index test? Unclear

**Could the conduct or the interpretation of the reference standard have introduced bias? Risk: Unclear**

**B. Applicability concerns**

**Are there concerns that the conduct or the interpretation of the reference standard do not match the review question? CONCERN: Low**

**DOMAIN 4: FLOW AND TIMING**

**A. Risk of Bias**

*Describe any patients who did not receive the index test and/or reference standard or who were excluded from the 2×2 table (refer to flow diagram)*: None.

*Describe the time interval and any interventions between index test and reference standard:* Unclear

- Was there an appropriate interval between index test and reference standard? Unclear
- Did all patients receive the same reference standard? Yes
- Were all patients included in the analysis? Yes

**Could the flow and timing have introduced bias? Risk: Unclear**

**QUADAS-2 Risk of bias and Applicability concerns Study Reference: Shin 2017**

**DOMAIN 1: PATIENT SELECTION**

**A. Risk of Bias**

*Describe methods of patient selection*: Patients with an indication for CAG due to chest pain or suspected CHD who were older than 18 years and suited for stress testing with MCG. Exclusion criteria: acute coronary syndromes or recent (b3 months) acute myocardial infarction, coronary artery bypass grafting, chronic total coronary occlusion, significant valvular heart disease, end stage renal failure, or refusal to enter the registry.

- Was a consecutive or random sample of patients enrolled? Unclear
- Was a case-control design avoided? Yes
- Did the study avoid inappropriate exclusions? Unclear

**Could the selection of patients have introduced bias? Risk: Unclear**

**B. Applicability concerns**

*Describe included patients (prior testing, presentation, intended use of index test and setting):* 96 subjects in total, including 75 males and 21 females. The subjects aged from 34 to 82 years, with a mean value of 65.0 ± 10.8 years.

**Are there concerns that the included patients and setting do not match the review question? CONCERN: Low**

**DOMAIN 2: INDEX TESTS**

**A. Risk of Bias**

*Describe the index test and how it was conducted and interpreted:* Performed using a 64-channel gradiometer system in a magnetically shielded room. The subject were in the supine position to record the rest MCG. Stress recordings were obtained by bicycle exercise test. Each signal is recorded for 100 s at a sampling rate of 500 Hz. An independent investigator performed quality evaluation and analysis of ECG and MCG. Patients with non-dipole phenomenon, which means the number of existed poles was either 1 or > 2, were diagnosed as CHD. For the inter-observer agreement of ischemic analysis by MCG (for ST-segment fluctuation score as well as non-dipole phenomenon analysis), kappa statistic was used.

- Were the index test results interpreted without knowledge of the results of the reference standard? Yes
- If a threshold was used, was it pre-specified? No

**Could the conduct or interpretation of the index test have introduced bias? Risk: High**

**B. Applicability concerns**

**Are there concerns that the conduce or interpretation of the Index test do not match the review question? CONCERN: Low**

**DOMAIN 3: REFERENCE STANDARD**

**A. Risk of Bias**

*Describe the reference standard and how it was conducted and interpreted*: CAG

*Readers:* Unclear

*Criteria for interpretation:* At least 70% stenosis in at least one proximal epicardial coronary artery and objective evidence of myocardial ischemia (substantial changes in ST-segment depression or T-wave inversion on the resting electrocardiogram or inducible ischemia with either exercise) or at least one coronary stenosis of at least 80% and classic angina without provocative testing.

- Is the reference standard likely to correctly classify the target condition? Yes
- Were the reference standard results interpreted without knowledge of the results of the index test? Unclear

**Could the conduct or the interpretation of the reference standard have introduced bias? Risk: Unclear**

**B. Applicability concerns**

**Are there concerns that the conduct or the interpretation of the reference standard do not match the review question? CONCERN: Low**

**DOMAIN 4: FLOW AND TIMING**

**A. Risk of Bias**

*Describe any patients who did not receive the index test and/or reference standard or who were excluded from the 2×2 table (refer to flow diagram)*: None.

*Describe the time interval and any interventions between index test and reference standard:* Within 24 hours.

- Was there an appropriate interval between index test and reference standard? Yes
- Did all patients receive the same reference standard? Yes
- Were all patients included in the analysis? Yes

**Could the flow and timing have introduced bias? Risk: Low**

**QUADAS-2 Risk of bias and Applicability concerns Study Reference: Zhao 2019**

**DOMAIN 1: PATIENT SELECTION**

**A. Risk of Bias**

*Describe methods of patient selection*: Patients who are pathologically diagnosed with MCG, radionuclide myocardial perfusion imaging (MPI), and CAG. Exclusion criteria: serious hypertension (blood pressure > 180/110 mmHg) OR complex arrhythmias (frequent ventricular premature contraction, ventricular tachycardia and complete bundle branch block) OR serious pulmonary disease and chest malformation or surgery OR < 3 months post-AMI OR ventricular hypertrophy or dilated cardiomyopathy confirmed by Echo OR valvular heart disease OR congenital cardiovascular disease OR post pacemaker OR heart failure NYHA > class III OR dysfunction of liver and kidney as well as abnormal electrolyte. All the patients signed written informed consent and are willing to cooperate with researchers.

- Was a consecutive or random sample of patients enrolled? Yes
- Was a case-control design avoided? Yes
- Did the study avoid inappropriate exclusions? Yes

**Could the selection of patients have introduced bias? Risk: Low**

**B. Applicability concerns**

*Describe included patients (prior testing, presentation, intended use of index test and setting):* 71 males and 33 females in total. Aged 58.56±8.13 and 54.74±8.58 years for CHD group and non-CHD group, respectively. No significant difference between groups for height and weight.

**Are there concerns that the included patients and setting do not match the review question? CONCERN: Low**

**DOMAIN 2: INDEX TESTS**

**A. Risk of Bias**

*Describe the index test and how it was conducted and interpreted:* Performed using a 9-channel mapping system in an unshielded room. The 9 sensors are arranged in a 3 × 3 rectangular grid and separated from each other by 4 cm. All magnetic, electronic and metallic objects were removed. MCG were recorded at four pre-defined positions for a total imaging time of 6 min. Surface ECG was recorded simultaneously as a reference signal for MCG signal averaging. The MCG data were analyzed in scalar form within T waves. System automatically drew an iso-magnetic map from 36 points of magnetic field component. With this map, 7 quantitative parameters were then calculated. The patients is diagnosed as CHD if at least 2 out of the following 7 indicators were found to be abnormal: 1) Pre-peak repolarization angle (PA) < -120 or > -25 2) Pre-peak repolarization trajectory (PT) ≥ 4.3 3) Pre-peak repolarization angular deviation (PAD) ≥ 0.5 4)Post-peak repolarization angle (PoA) < -110 or > -22 5) Post-peak repolarization trajectory (PoT) ≥ 3.7 6) Post-peak repolarization angular deviation (PoAD) ≥ 0.45 7) Pre-Post angle change (PPAC) < -35 or > -12

- Were the index test results interpreted without knowledge of the results of the reference standard? Unclear
- If a threshold was used, was it pre-specified? Unclear

**Could the conduct or interpretation of the index test have introduced bias? Risk: Unclear**

**B. Applicability concerns**

**Are there concerns that the conduce or interpretation of the**

**Index test do not match the review question?** **CONCERN: Low**

**DOMAIN 3: REFERENCE STANDARD**

**A. Risk of Bias**

*Describe the reference standard and how it was conducted and interpreted*: CAG

*Readers:* Unclear

*Criteria for interpretation:* Narrowing of the coronary arteries >= 50% in one or more vessels.

- Is the reference standard likely to correctly classify the target condition? Yes
- Were the reference standard results interpreted without knowledge of the results of the index test? Unclear

**Could the conduct or the interpretation of the reference standard have introduced bias?** **Risk: Unclear**

**B. Applicability concerns**

**Are there concerns that the conduct or the interpretation of the reference standard do not match the review question?** **CONCERN: Low**

**DOMAIN 4: FLOW AND TIMING**

**A. Risk of Bias**

*Describe any patients who did not receive the index test and/or reference standard or who were excluded from the 2×2 table (refer to flow diagram)*: None.

*Describe the time interval and any interventions between index test and reference standard:* Unclear

- Was there an appropriate interval between index test and reference standard? Unclear
- Did all patients receive the same reference standard? Yes
- Were all patients included in the analysis? Yes

**Could the flow and timing have introduced bias?** **Risk: Unclear**

**QUADAS-2 Risk of bias and Applicability concerns** **Study Reference: Shin 2019**

**DOMAIN 1: PATIENT SELECTION**

**A. Risk of Bias**

*Describe methods of patient selection*: Consecutive patients older than 18 years and suited for stress testing with MCG, who were admitted to the hospital with an indication for CAG due to chest pain or suspected CHD. Exclusion criteria: acute coronary syndromes, recent (<3 months) acute myocardial infarction, coronary artery bypass grafting, chronic total coronary occlusion, significant valvular heart disease, end stage renal failure, or refusal to enter the registry.

- Was a consecutive or random sample of patients enrolled? Yes
- Was a case-control design avoided? Yes
- Did the study avoid inappropriate exclusions? Unclear

**Could the selection of patients have introduced bias? Risk: Unclear**

**B. Applicability concerns**

*Describe included patients (prior testing, presentation, intended use of index test and setting):* See above. 129 males and 73 females in total, with a mean age of 64.6 years.

**Are there concerns that the included patients and setting do not match the review question? CONCERN: Low**

**DOMAIN 2: INDEX TESTS**

**A. Risk of Bias**

*Describe the index test and how it was conducted and interpreted:* Performed using a 64-channel gradiometer system in a magnetically shielded room. The rest MCG were recorded for 100 seconds at a sampling rate of 500 Hz, with the patient in the supine position and the SQUID's 2-D arrayed sensors positioned close to, but not in contact with the left chest wall. Stress recordings were acquired by bicycle exercise test. One independent investigator performed quality evaluation and analysis of ECG and MCG. The patients with the percent change of ST–segment fluctuation score ≥ −40.0% is diagnosed as CHD.

- Were the index test results interpreted without knowledge of the results of the reference standard? Unclear
- If a threshold was used, was it pre-specified? No

**Could the conduct or interpretation of the index test have introduced bias? Risk: High**

**B. Applicability concerns**

**Are there concerns that the conduce or interpretation of the Index test do not match the review question? CONCERN: Low**

**DOMAIN 3: REFERENCE STANDARD**

**A. Risk of Bias**

*Describe the reference standard and how it was conducted and interpreted*: CAG

*Readers:* Unclear

*Criteria for interpretation:* >= 70% luminal obstruction.

**Could the conduct or the interpretation of the reference standard have introduced bias? Risk: Unclear**

**B. Applicability concerns**

**Are there concerns that the conduct or the interpretation of the reference standard do not match the review question? CONCERN: Low**

**DOMAIN 4: FLOW AND TIMING**

**A. Risk of Bias**

*Describe any patients who did not receive the index test and/or reference standard or who were excluded from the 2×2 table (refer to flow diagram)*: None.

*Describe the time interval and any interventions between index test and reference standard:* Unclear

- Was there an appropriate interval between index test and reference standard? Unclear
- Did all patients receive the same reference standard? Yes
- Were all patients included in the analysis? Yes

**Could the flow and timing have introduced bias? Risk: Unclear**

**QUADAS-2 Risk of bias and Applicability concerns** **Study Reference: Huang 2019**

### DOMAIN 1: PATIENT SELECTION

#### A. Risk of Bias

*Describe methods of patient selection*: Consecutive patients with suspected CHD exhibiting chest pain or chest discomfort. Exclusion criteria included: 1. Malignant tumor. 2. Structural heart disease 3. Valvular heart disease 4. Cardiomyopathy. 5. Malignant arrhythmias (atrial fibrillation, atrial flutter, and ventricular arrhythmias).

- Was a consecutive or random sample of patients enrolled? Yes
- Was a case-control design avoided? Yes
- Did the study avoid inappropriate exclusions? Unclear

**Could the selection of patients have introduced bias? Risk: Unclear**

#### B. Applicability concerns

*Describe included patients (prior testing, presentation, intended use of index test and setting):* For CHD group, the mean value of age is 59.7 ± 11.3 years. There are 102 males in the total of 132 CHD patients. For non-CHD group, the mean value of age is 59.5 ± 9.2 years. There are 38 males in the total of 81 non-CHD patients.

**Are there concerns that the included patients and setting do not match the review question? CONCERN: Low**

### DOMAIN 2: INDEX TESTS

#### A. Risk of Bias

*Describe the index test and how it was conducted and interpreted:* Performed using 4-channel MCG system in an unshielded room. All magnetic, electronic and metallic objects were removed. The sensors were placed at 1-2 cm above the chest and recorded 36 points covering a rectangle area of 25 × 25 cm. These signals were then sent to the computer and analyzed. The patient is diagnosed as CHD if at least 1 out of the following 4 indicators were found to be abnormal: the angel of T peak magnetic field > 12.205, the change of the angle of TT current > 62.625, the change of the angle of TT > 43.215, the minimux of the angle of TT current < -27.725.

- Were the index test results interpreted without knowledge of the results of the reference standard? Unclear
- If a threshold was used, was it pre-specified? No

**Could the conduct or interpretation of the index test have introduced bias? Risk: High**

#### B. Applicability concerns

**Are there concerns that the conduce or interpretation of the Index test do not match the review question? CONCERN: Low**

### DOMAIN 3: REFERENCE STANDARD

#### A. Risk of Bias

*Describe the reference standard and how it was conducted and interpreted*: CAG

*Readers:* Unclear

*Criteria for interpretation:* At least one coronary artery stenosis >= 50% in the three main coronary arteries and their branches.

- Is the reference standard likely to correctly classify the target condition? Yes
- Were the reference standard results interpreted without knowledge of the results of the index test? Unclear

**Could the conduct or the interpretation of the reference standard have introduced bias? Risk: Unclear**

**B. Applicability concerns**

**Are there concerns that the conduct or the interpretation of the reference standard do not match the review question? CONCERN: Low**

---

**DOMAIN 4: FLOW AND TIMING**

**A. Risk of Bias**

*Describe any patients who did not receive the index test and/or reference standard or who were excluded from the 2×2 table (refer to flow diagram)*: None.

*Describe the time interval and any interventions between index test and reference standard:* Unclear.

- Was there an appropriate interval between index test and reference standard? Unclear
- Did all patients receive the same reference standard? Yes
- Were all patients included in the analysis? Yes

**Could the flow and timing have introduced bias? Risk: Unclear**

**QUADAS-2 Risk of bias and Applicability concerns** **Study Reference: Coriasso 2021**

**DOMAIN 1: PATIENT SELECTION**

**A. Risk of Bias**

*Describe methods of patient selection*: Patients with suspected CHD. Exclusion criteria and recruitment procedure unclear.

- Was a consecutive or random sample of patients enrolled? Unclear
- Was a case-control design avoided? Yes
- Did the study avoid inappropriate exclusions? Unclear

**Could the selection of patients have introduced bias? Risk: Unclear**

**B. Applicability concerns**

*Describe included patients (prior testing, presentation, intended use of index test and setting):* 62% male. Mean age of 60.9 years old. Inclusion criteria see above.

**Are there concerns that the included patients and setting do not match the review question? CONCERN: Low**

**DOMAIN 2: INDEX TESTS**

**A. Risk of Bias**

*Describe the index test and how it was conducted and interpreted:* Patients whose MCG displays the presence of "RT angle", "multipolar", or "island" is diagnosed as CHD. Procedure for clinical examination and data collection not reported.

- Were the index test results interpreted without knowledge of the results of the reference standard? Yes
- If a threshold was used, was it pre-specified? Unclear

**Could the conduct or interpretation of the index test have introduced bias? Risk: Unclear**

**B. Applicability concerns**

**Are there concerns that the conduce or interpretation of the Index test do not match the review question? CONCERN: Low**

**DOMAIN 3: REFERENCE STANDARD**

**A. Risk of Bias**

*Describe the reference standard and how it was conducted and interpreted*: CAG

*Readers:* Unclear

*Criteria for interpretation:* Unclear.

- Is the reference standard likely to correctly classify the target condition? Yes
- Were the reference standard results interpreted without knowledge of the results of the index test? Unclear

**Could the conduct or the interpretation of the reference standard have introduced bias? Risk: Unclear**

**B. Applicability concerns**

**Are there concerns that the conduct or the interpretation**

**of the reference standard do not match the review question?** **CONCERN: Low**

**DOMAIN 4: FLOW AND TIMING**

**A. Risk of Bias**

*Describe any patients who did not receive the index test and/or reference standard or who were excluded from the 2×2 table (refer to flow diagram)*: None.
*Describe the time interval and any interventions between index test and reference standard:* Unclear.

- Was there an appropriate interval between index test and reference standard? Unclear
- Did all patients receive the same reference standard? Yes
- Were all patients included in the analysis? Yes

**Could the flow and timing have introduced bias?** **Risk: Unclear**
